# Tensor cardiography: a novel ECG analysis of deviations in collective myocardial Action Potential transitions based on point processes and cumulative distribution functions

**DOI:** 10.1101/2023.05.13.23289858

**Authors:** Shingo Tsukada, Yu-ki Iwasaki, Yayoi Tetsuo Tsukada

**Affiliations:** NTT Basic Research Laboratories, Bio-Medical Informatics Research Center, 3-1, Morinosato Wakamiya, Atsugi-city, Kanagawa Pref., 243-0198 Japan Premium Research Institute for Human Metaverse Medicine (WPI-PRIMe), Osaka University; Department of Cardiovascular Medicine, Nippon Medical School; Department of General Medicine and Health Science, Nippon Medical School

## Abstract

A method to estimate myocardial action potentials (APs) from electrocardiograms (ECGs) would be an advance in ECG-based diagnosis, utilised for clinical diagnosis, assessment of potential cardiac disease risk and prediction of lethal arrhythmias. However, the ECG inverse problem, which estimates the spatial distribution of AP signals from the ECG, has been considered difficult electromagnetically. For clinical ECG analysis, timescales of collective APs, synchrony and the duration of depolarisation and repolarisation is informative. Thus, we attempted to obtain the time distribution of collective AP transitions from the ECG rather than the spatial distribution.

To analyse the variance of the collective myocardial APs from the ECG, we designed a model equation using the probability densities of the Gaussian function of time-series point processes in the cardiac cycle and dipoles of collective APs in the myocardium. The equation to calculate the difference between the two cumulative distribution functions (CDFs) as the positive- and negative-epicardium potential fits well with the R and T waves. The mean, standard deviation, weights, and level of each CDFs are metrics for the variance of the AP transition state of the collective myocardial AP transition states. Clinical ECGs of myocardial ischaemia during coronary intervention showed abnormalities in the aforementioned specific elements of the tensor associated with repolarisation transition variance earlier than in conventional indicators of ischaemia. The tensor could evaluate the beat-to-beat dynamic repolarisation changes between the ventricular epi and endocardium using the Mahalanobis distance (MD). Tensor Cardiography, a method that uses CDF differences CDF as the transition of a collective myocardial AP transition, has the potential to be a new analysis tool for ECGs.

**Author’s Summary:** Myocardial action potentials (APs) which indicate electric excitation of the cells can provide important information to suggest the mechanisms of cardiac disease such as myocardial ischemia and arrhythmias. However, it has been challenging to estimate APs from electrocardiograms (ECGs). Unlike other imaging techniques like CT or MRI, the electrocardiographic inverse problem requires estimating the geometric distribution of APs from the ECG, has been considered difficult.

Our approach, known as Tensor Cardiography, uses a model equation based on cumulative distribution functions (CDFs) to analyze the time series variance of collective myocardial APs from the ECG. By fitting this equation to the R and T waves, we have obtained a set of metrics that represent beat-to-beat dynamic variance of polarization and repolarization of the epi and endocardium. Our study of ECGs from myocardial ischemia during coronary intervention has demonstrated abnormalities in the tensor elements associated with repolarization, which appeared earlier and more prominently than conventional ST changes.

Tensor Cardiography provides a revolutionary analysis tool for ECGs that holds enormous potential for clinical diagnosis, risk assessment, and prediction of lethal arrhythmias. Our approach shows promise as a new frontier in cardiac disease management and has significant implications for patient care.

## Introduction

Developed by Willem Einthoven in 1901, the electrocardiogram (ECG) has been widely used in clinical practice to this day as a simple, inexpensive, non-invasive tool for evaluating the heart’s electrical phenomena.(1) (2)However, ECG diagnosis is quite difficult even for cardiologists because of the broad range of normal variance. ECG analysis is conducted in accordance with restricted guidelines such as voltage, width, potential and interval of the P-QRS-T wave, electrical axis and ST segment deviation(3).(4) In addition, it is difficult to clearly evaluate the minute distortions on the border between a normal and abnormal ECG, which are known as nonspecific changes related to potential heart diseases.

Various efforts have been made to detect abnormalities and identify ECG waveforms. Machine learning, e.g., deep learning methods such as continuous recurrent neural networks or long short-term memory, has also been used(5). Deep learning of ECGs reveals that subtle distortions of ECG waveforms contain unknown features and invisible latent information related to previously unnoticed heart diseases and conditions. However, these features are often difficult to explain and use as quantitative metrics.

A statistical analysis of the variations in collective myocardial APs based on ECGs would be useful in understanding the pathophysiology and diagnosis of the disease.

A method to estimate myocardial action potentials (APs) from electrocardiograms (ECGs) would be an advance in ECG-based diagnosis, utilised for clinical diagnos is, assessment of potential cardiac disease risk and prediction of lethal arrhythmias. However, contrary to inverse problems such as computerized tomography (CT) from X-ray radiographs, and magnetic resonance imaging (MRI) from nuclear magnetic resonance, electrocardiographic inverse problem, which estimates the geometric distribution of APs from the ECG, has been considered difficult.

Despite the limitations of its geometric representation, the dipole model is widely used to explain the time axis relationship of APs and ECG. For example, the dipole concept is used to interpret the characteristic Coved-type electrocardiogram of Brugada syndrome, and is used as a reasonable model clinically(6).

For time series analysis of myocardial action potentials and the ECG, it is known that the action potential of the endocardial myocardium (anode) and the action potential of the epicardial myocardium (cathode) differences based on the dipole are approximately equal to the ECG. We modelled the dipole concept using a probability density function and designed a method to estimate the anode and cathode components of depolarization and repolarization separately from the ECG.

Specifically, the difference between the two cumulative distribution functions two CDFs fits with the R and T waves separately by least squares method show a high representativeness of the ECG and obtain the metrics of 4 CDF’s mean, standard deviation, weights, level and intervals, which form fourth order tensor by combining with the time series of beats and the number of ECG lead channels. The graph and the metrics of four CDFs tensor indicate the variance of collective myocardial anodal and cathodal APs separately, which are hereafter referred to as “Tensor Cardiography (TCG)”.

## Methods

### Modelling: relation between ECG and myocardial APs

Previous studies have shown that the relationship between ventricular cardiomyocyte APs and ECGs is mainly formed by the asymmetric structure of the ventricular muscle and the non-uniform distribution of myocardial potentials caused by AP propagation patterns of APs originating from the ventricular endocardium leached to the epicardium, the basal to the apex (7) (Figure 1A). APs on the endocardial side of the ventricle (including middle layer, M-cell(8) (9)) have a longer duration and give positive potentials to the ECG II leads, while those on the epicardial side have a shorter duration and give negative potentials to the ECG. The ECG corresponds approximately to the difference between the AP of the endocardial side minus that of the epicardial side(10) (Figure 1B).

**Figure 1.**
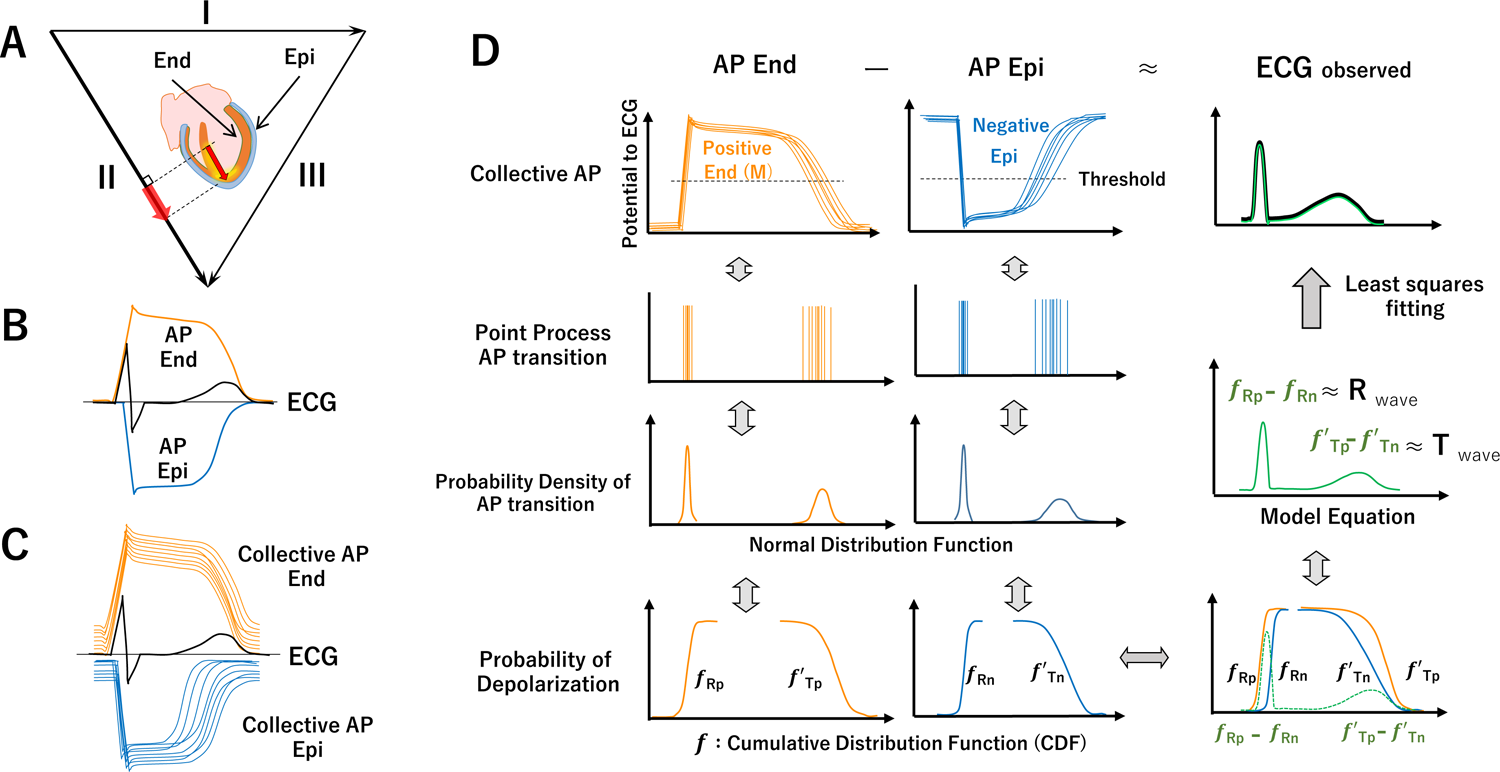
**A. B** Asymmetry of ventricular wall and AP propagating from endocardial side to epicardial side and repolarisation from Epi to End (M cell) generating R and T waves of ECG (lead II). **C,** AP collective shows the variance of time axis. **D,** Endo and Epi AP elicit positive and negative potential, respectively. Their difference is closely related to ECG. Timing of AP transition (threshold crossisng) represented by point process, probability density (normal distributions) and probability (cumulative distribution functions, CDFs *f_Rp_ f’_Rn_ f’_Tp_ f’_Tn_)*. Relation of End AP (f _Rp_ f’_Tp_,) and Epi Ap, f’_Rn_ f’_Tp_ and the difference of depolarisation (f_Rp_-f’_Rn_,), and repolarisation (*f_Tp_-f’_T_*_n_) approximate to ECG R and T waves, respectively.

### ECG probabilistic model

The distribution of collective ventricular muscle AP transitions (the timing of the membrane potential threshold crossing) (Figure 1C) is modelled by a time-series point process (Figure 1D). The timing of the transitions is synchronised by the conduction system and has the physiological variability; depolarization points are relatively densely distributed and repolarisation points are sparsely distributed. A normal distribution is used as the function for this probability density distribution since a myocardium collective is large and commonly used for biomedical statistical modelling.

### Cumulative distribution function (CDF) for collective myocardial AP transitions

Ventricular muscles maintain an AP for a certain duration. Thus, the probability of a ventricular muscle from phase 0 (1) to phase 2 (ON state) can be expressed by a cumulative distribution function (Figure 1D, CDF, Supplement Figure 1). In contrast to depolarisation, the repolarisation state (phase 2 to phase 3 OFF state) is a process in which the probability of a ventricular muscle in the ON state progressively decreases over time. For the repolarisation transition process, the CDF is in the opposite direction to the time ax is decreasing from 1, hereinafter referred to as the “inverse cumulative distribution function (inverse CDF)” with greater variability than the depolarisation.

### Difference of CDF, equation of ECG for AP transition approximation

#### ECG modelling by bipolar CDFs

The relationship between the ECG and a cardiomyocyte AP is a very complex system(11) (12). To describe complex phenomena simply, the ECG is modelled using a polarity-marked point process in which the AP exerts a positive (anodic) or negative (cathodic) binary potential change on the ECG and the sum of them corresponds to the ECG. Specifically, the collective myocardium that contributes to the ECG of a specific lead (e.g., induction II) consists of a group of myocardium that produces an anodic electromotive force (EMF) to the ECG during the ON/OFF transition and a group of myocardium that produces a cathodic EMF.

Since the binary potential distribution of the anodic and cathodic groups APs are combined with opposing polarities, the difference between them can be assumed to be approximately equivalent to the ECG (13) (14). In other words, the ECG is probabilistically represented by approximating the difference between the transition processes of the anodic and cathodic groups by a positive - negative (anodic - cathodic) signed point process to the ECG. More specifically, the collective myocardium AP transition process is represented in the probability density function using four CDFs, two for each process of depolarisation and repolarisation (Figure 2A).

**Figure 2.**
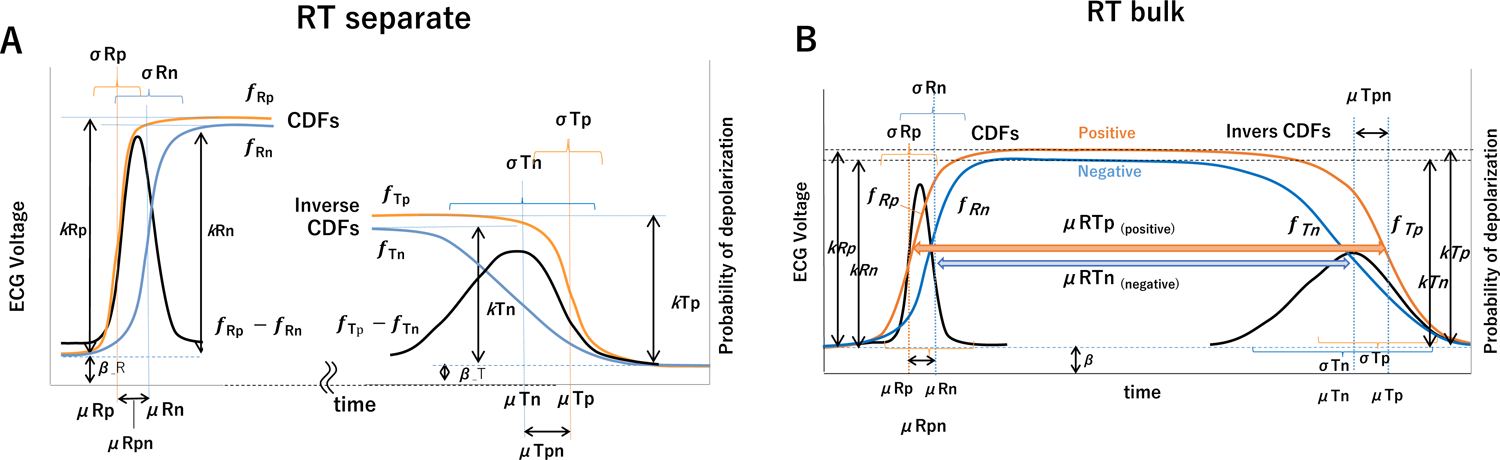
**A**, RT separation: parameters of TCG when R and T are calculated separately **B,** RT Bulk: Parameters of TCG for RT integrated calculation under the condition that APs are combined at plateau.

The transition process of R wave from a resting membrane potential to depolarisation is represented by an anodic and cathodic cumulative probability function (fRp and fRn, respectively). For ECG II lead, due to the asymmetry of the ventricle, the AP of the inner (endocardial) outer (epithelial) sides of the ventricular wall contribute to the anodal and cathodal potential, respectively. According to the order of excitation, depolarisation starts from the inner (anode) side of the ventricular wall and propagates to the outer (cathode) side, and the difference between the two sides of the CDF (fRp – fRn) corresponds to the ECG R wave.

The T wave is the process of transitioning from a depolarised state to a resting membrane potential. Since depolarisation and repolarisation are opposite in polarity, and the propagation of repolarisation is in the reverse order of depolarisation due to the difference in the APD of the ventricular layer structure and function, the difference between the inner (anode) and outer (cathode) inverse CDFs (*f’Tp – f’Tn*) corresponds to the ECG.

The difference between the measured ECG and this equation can be minimised by the least squares method. The two CDFs (above *fR_p_ fR_n_*) and two inverse CDFs (*f’T_n_ f’T_p_*) obtained by ECG approximation represent the AP transition process time series of the anode and cathode groups of the origin dipole of the target R and T waveforms, respectively.

Although the CDFs represent probabilities from base 0 to maximum 1, the four CDFs obtained by fitting the ECGs often result in different heights of maximum 1 (probability). Each height (weight) reveals information about the ECG R- and T-wave potentials and the combined relative anodic and cathodic potentials of the waves.

In this processing, two CDFs difference equations were fitted independently to the QRS interval of the R wave and to the interval from the start to end of the T wave were independently fitted with the difference equations of the two CDFs. Thus this way referred to as “RT separate” method (Figure 2A).

Since depolarisation and repolarisation are linked with each cardiac contraction, the CDF and reverse CDF pairs for each anode (*fR_p_-f’T_p_*) and cathode (*fR_n_ -f’T_n_*) can be connected in plateau sections by using the least squares method with the restriction that the CDF and inverse CDF are connected at the plateau (Figure 2B), To horizontally connect the anodic CDF f_Rp_ and in verse CDF f_Tp_, and the cathodic CDF fRn and inverse CDF f_Tn_, respectively, constraints were applied to equalize the T-wave height by the weights k_Tp_ and k_Tn_.

As a result, the probability densities of depolarisation are expressed as two trapezoids with the left leg being steeper and the right leg being slower. The horizontal level of each plateau represents the probability equal to 1 and the level of baseline represents 0 (Supplement Figure 1, Depolarised Probability Graph).

Converting the four CDFs into probability density functions (unimodal normal distribution) clearly shows the frequency when the APs of the depolarisingand repolarising anode and cathode groups transition in the time series, which were referred to as “RT Bulk” method (Figure 2B).

### ECG Data from healthy participants

Physio net Open ECG Data of a healthy participants, “Autonomic Aging” and “MIT-BIH ST change” database was used for the standard value of TCG(15).(16)

### Clinical ECG data

We apply two clinical ECG datasets: patients with effort angina and early repolarisation syndrome. These data were obtained from the Department of Cardiovascular Medicine, Nippon Medical School.

### Statistical Analysis

ANOVA with Tukey honestly significant difference (HSD) and correlation coefficient was used for statistical processing. The p-values lower than 0.05 were considered significant. The Mahalanobis distance (MD) was used as a multivariate distance measure to identify similarities and differences between a single heartbeat and multiple heartbeats.

### Data Availability

We are currently consulting with the Ethics Committee of Nippon Medical School regarding the disclosure of ECG and TCG data included in the case reports (PCI and ERS. If approved, the corresponding author will provide the data upon request.

## Results

The application of TCGA to the II-lead ECG of a healthy participant is shown in Figure 3. The P-QRS-T point was obtained from the inflection point and standard time interval (Figure 3, red points).

**Figure 3.**
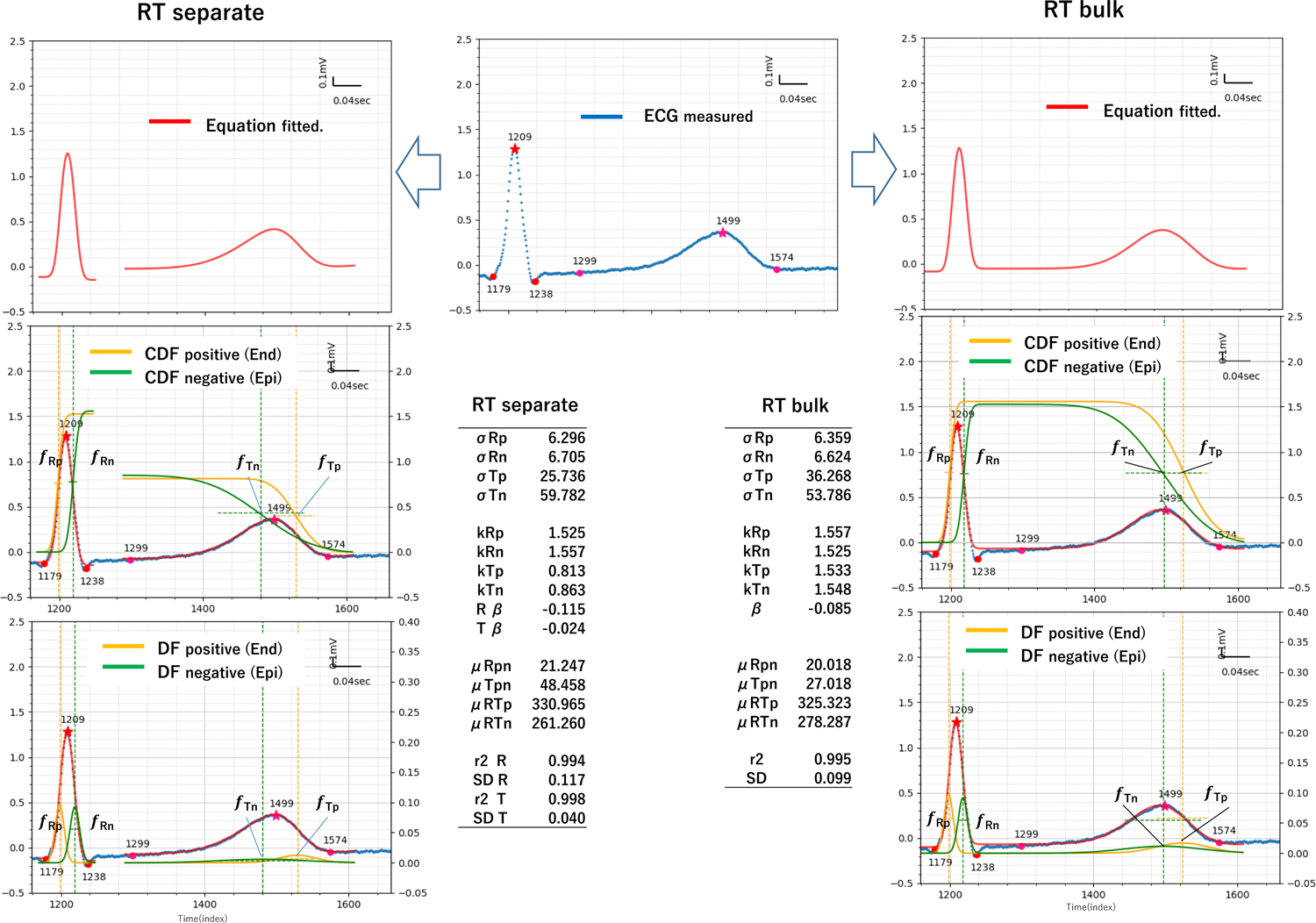
Example of TCG RT separation (left) RT Bulk (right), blue dotted line: observed (target) ECG (middle), red line: TCG equation fitted to ECG minimised least square method, orange line: positive (Endo, green line: negative (Epi). The bottom box shows the result of the TCG parameters.

### RT separate method

In the RT separate method, two CDFs difference equations were fitted independently to the QRS interval of the R wave and to the interval from the start to end of the T wave (Figure 3, dotted blue line) were independently fitted with the difference equations of the two CDFs.

Specifically, the difference equations fRp - fRn and fTp - fTn of the two CDFs were approximated to the QRS interval of the R wave and the T-wave interval (from the start, peak and end point of the T wave), respectively, using the least squares method. The anodic CDF fRp and inverse CDF fTp obtained from the fitting are represented by the orange lines, and the cathodic CDF fRn and inverse CDF fTn obtained from the fitting are represented by the green line s. The results of the approximation equation are shown by the red line. Smooth waveforms were obtained without noise found in the original waveforms.

The coefficient of determination (r^2^) of the QRS and T waves are 0.994 and 0.998, respectively (QRS SD= 0.117, T SD=0.040).

The resultant four CDFs (fRp, fRn, fTp, fTn), mean (μRp, μRn, μTp, and μTn), standard deviation (σRp, σRn, σTp, σTn), weight (kRp,kRn,kTp,kTn) and level (βR, βT) were obtained (Figure 3 left lower Table).

The equation for the CDF difference yields two CDFs each from the R and T waves, and in most cases, the original waveforms are fitted with a coefficient of determination of 0.95 or higher, yielding good fitting results. However, if the quality of the ECG signal was problematic, i.e., if the ECG contained distortion, base line drift or noise, the fitting may have failed or abnormal values were output.

To determine the standard values of each TCG parameter and the difference of age groups, Physionet “Autonomic Aging” data were examined. Table 1A shows the results of TCG performed on 699 participants selected from the ECG data of Physionet Autonomic Aging, which were recorded clearly with less noise. TCG parameters were calculated for 500 heartbeats per participant and the average value was calculated to be the result for each participant. The mean values and standard deviations for the 699 participants are also shown in the table1.

**Table 1.**
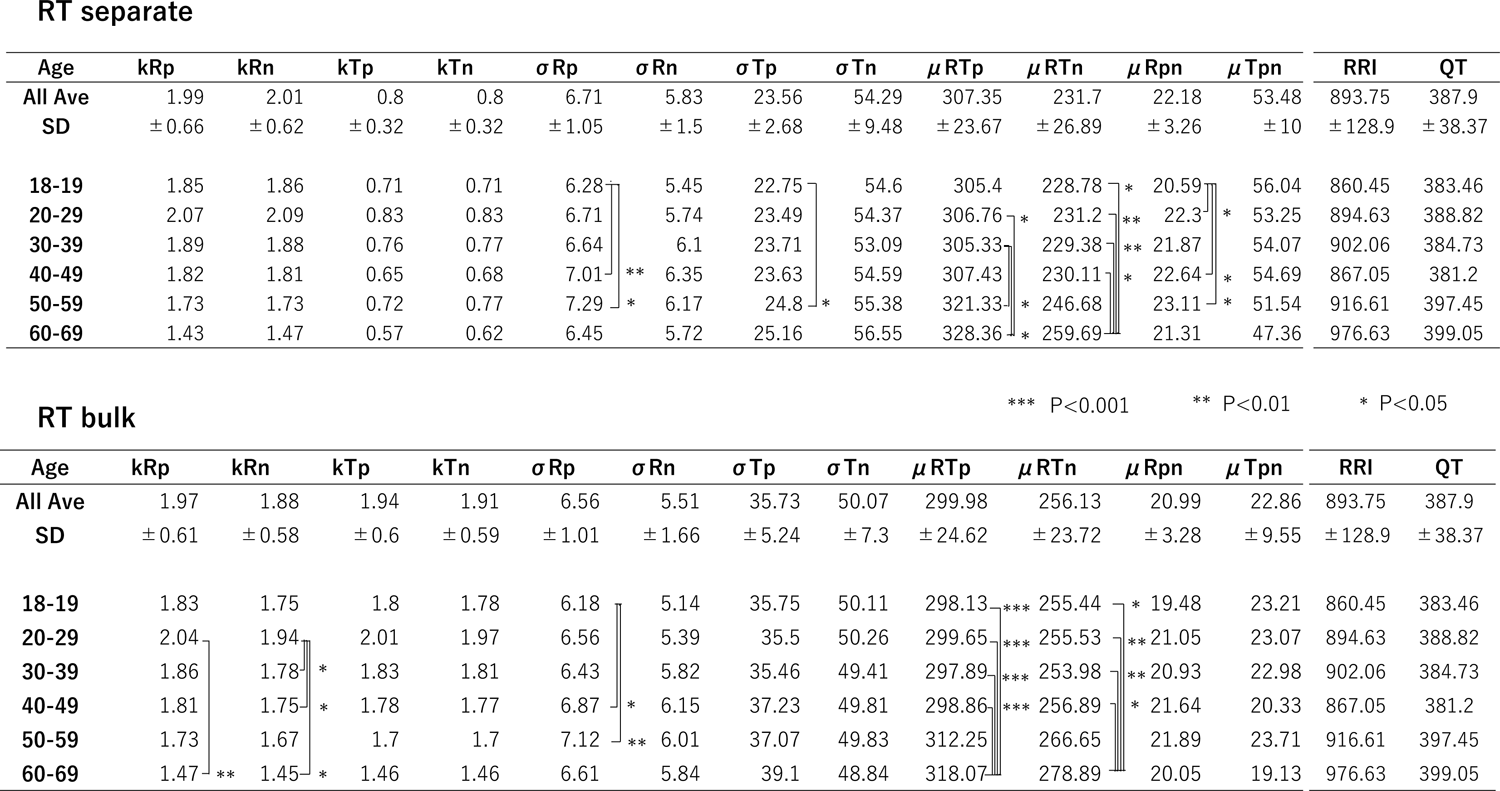

The time series is in the order of μRp, μRn, μTn, μTp. The standard deviations σRp, σRn, σTp, σTn are distributions (width of time series), and the relationship σRn<σRp<< σTp<σTn is observed. The heights kRp, kRn, kTp, kTn represent the relative weights of each function.

In terms of the effect of age on TCG and the correlation of each TCG parameter, the most of TCG parameters were stable and showed little change with age. Contrary, significant differences in time-related TCG parameters, prolongation of μRTp, μRTn and decrease of μRTp and μRTn were observed in the 60-69y group (Table1).

### RT Bulk method

The RT separate method yields different heights of the four CDFs of TCG, and the CDF and inverse CDF are not horizontally connected. Therefore, to horizontally connect the anodic CDF fRp and inverse CDF fTp, and the cathodic CDF fRn and inverse CDF fTn, respectively, constraints were applied to equalise the T-wave height by the weights kTp and kTn (RT Bulk method, Figure 2B and Figure 3, right side).

The coefficient of determination (r^2^) of the Bulk method is 0.995 (Bulk SD=0.099). The RT separation and Bulk methods showed slightly different results for me an, standard deviation and weight (Figure 3 right side, Table 1B). The Bulk method tended to fit broadly in the time axis direction, including the ST level, while the separation method focused on the vertical amplitude direction of the R and T waves, and was believed to have been caused by the different fitting areas of the R, T and ST waves.

The correlation coefficients between TCG parameters with relation to time (interval) and R-R interval (RRI), QT are shown in Supplement Table 1A, and the correlation coefficients for TCG parameters related to potential are shown in Supplement Table 1B (the scatter plots of them are shown in Supplement Figure 2).

QT is correlated with μRTp μTnp, RRI is correlated with μRTp, and μRTp and μRTn are correlated. kRp and kRn are highly correlated with each other and with the height of the R wave. kTp and kTn are highly correlated with each other and with the height of the T wave. The ST level correlates with Tβ, and a correlation is also observed between Rβ and Tβ.

### Clinical application of TCG: case report presentation

#### Case 1: A male patient with angina underwent percutaneous coronary intervention (PCI) for left anterior descending artery (LAD)

We investigated the relationship between the ECG changes and TCG parameters during elective PCI for 90% stenosis at left anterior descending coronary artery (#6–7). This patient showed ST level changes with transient myocardial ischaemia induced by balloon inflation for three times. Figure 4A

1. In the resting ECG before balloon inflation, the ST level was normal (ECG V5 lead, blue dot line). The TCG (Bulk method) result shows that the heights of cathodic and anodic plateau level were almost equal. The cathodic k _p_f, (greenT Tp lines) represented the elongation of repolarisation, resulting in the kTp rTp and *k_Tn_ r_Tn_* crossing (black arrow).
2. During the left coronary angiography with contrast medium, the ECG showed ST depression and an increase of R waves indicating myocardial ischaemia. TCG showed fRn is markedly elevated than fRp. In addition, a backward shift of *kTp rTp* and *kT nrTn* was observed (orange arrow).
3. Transient myocardial ischaemia was induced by coronary artery occlusion by balloon inflation resulting in increase of T-wave amplitude on the ECG (red arrow); TCG showed the enlargement of the gap between fTp and fTn due to a forward shift of fTn (green arrow).
4. The second balloon inflation also elicited an increase of T waves on the ECG. TCG results indicate marked elongation of cathodal repolarisation (green line, downward slope).
5. The third balloon inflation resulted in a prolonged QT on the ECG; TCG showed a marked backward shift in fTn and fTp (orange and green arrows).
6. In the ECG after the completion of the PCI, the ST depression disappeared. TCG results show equal heights of fRp and fRn (fTp and fTn). The time direction shift of *rT* were also reduced, but the *kTp rTp* and *kT n rTn* crossing remains (black arrow).

**Figure 4.**
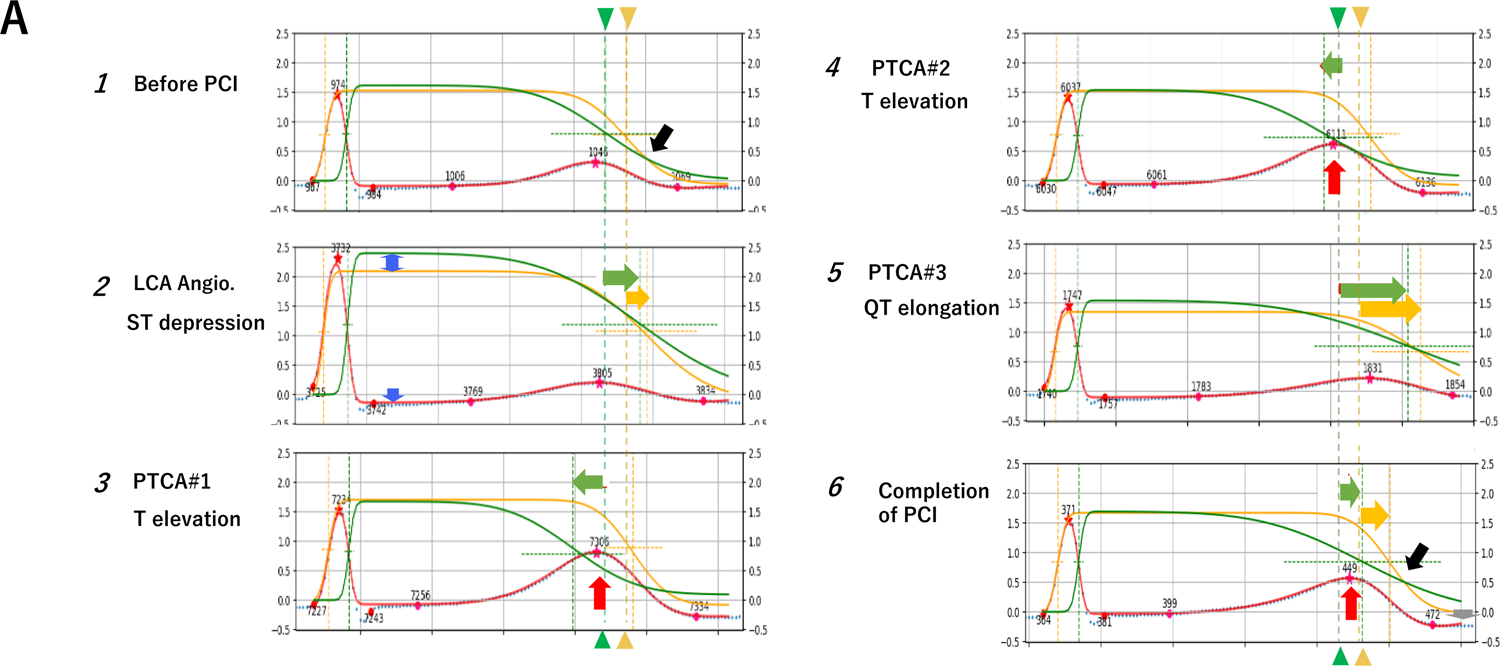
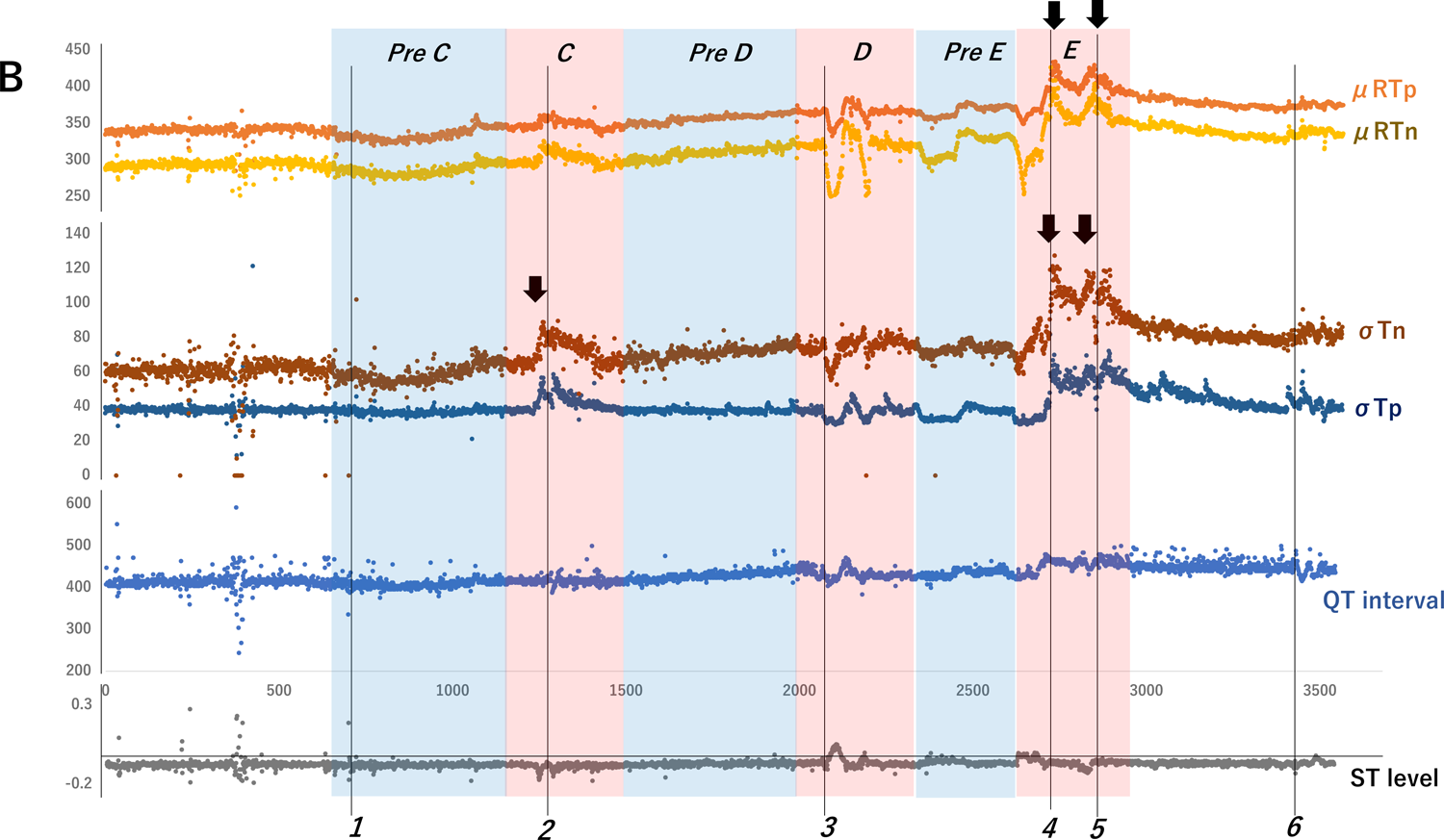
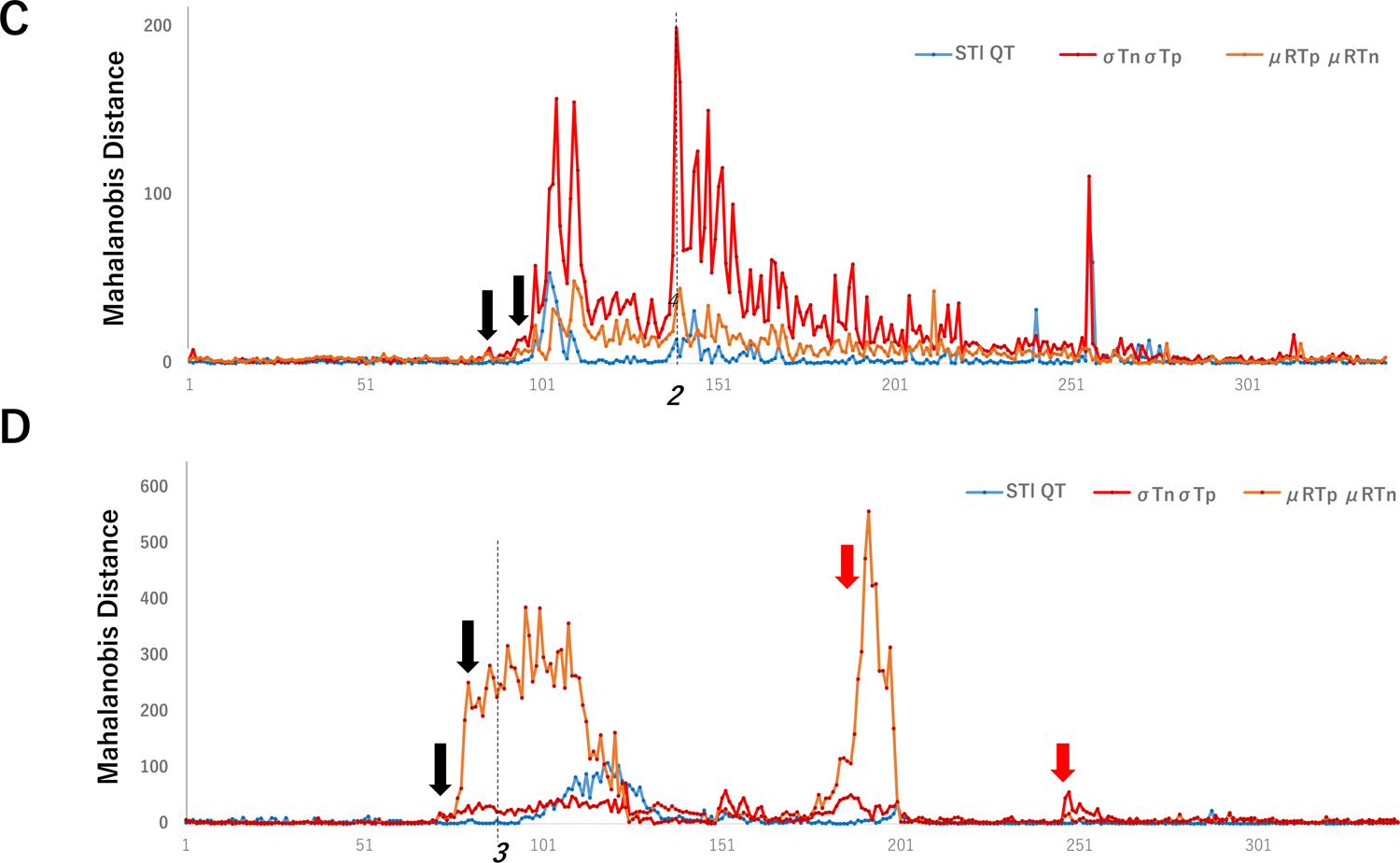
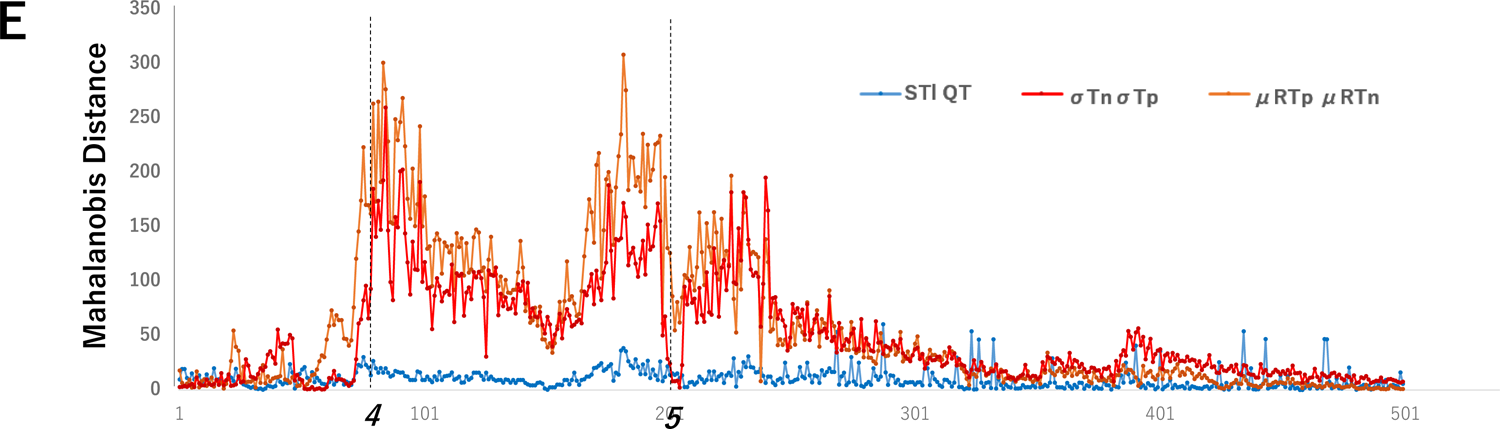
TCG parameters of patient with angina during elective PCI. **A,** Graphs of TCG (RT Bulk) equation to ECG during the intervention **1.** before PCI, **2.** LCA angiography: ST depression, **3.** First PTCA: T elevation, **4.** Second PTCA: T elevation **5.** Third PTCA: QT elongation, **6.** Completion of PCI: T elevation, (1∼6 locations indexed in **B**). Blue dotted line: Target ECG, Red line: fitting result of TCG equation to ECG, Orange line: positive (Endo) CDF, Green line: negative (Epi) CDF, vertical dotted cross line represents the location of the mean of CDF. Arrow heads marked original location (1) of the mean. Horizontal block arrows (Green and yellow) indicating repolarisation time shifts during PCI. Red arrow indicating T-wave elevation. Black arrows indicating crossing of Epi End inversion of repolarization. **B,** Time-series graph of TCG time-related parameters, μRTp, μRTn, σTp, σTn and the conventional ST level, QT interval during the intervention. The numbers at the bottom indicate times of TCG graphs in **A.** Black arrows indicate points that are not detected by conventional indicators but are detected by TCG parameters. **C,** Mahalanobis Distance (MD) of conventional indexes ST level and QT interval (blue line) and TCG parameters σTn andσT’(red line), μRTp and μRTn (orange line) between the intervals of *pre C* (reference) and *C* (target), marked by blue and red rectangles in **B** respectively. **D,** The MD between *pre D – D****_E_*** The MD between *pre E-E*

Figure 4B shows a time-series graph of the TCG time-related parameters μRTp, μRTn, σTp, σTn and the conventional ST level and QT interval during the intervention (Figure 4B). σTn,σTp, μRTn and μRTp showed markedly fluctuated and long-lasting responses (black arrows).

Table 2 showed the TCG parameter values at time points *1–6*. Bolded va lues indi cated notable values at each time point from A to F. ST depression (2) affected bulkσTp,σTn, μTpn. T-wave amplitude increased μTpn (*3, 4*) and kTp (*3*), QT elongation (*5*) elongatedσTn,σTp, μRTp, μRTn (Table 2).

**Table 2.**

| | kTp | kTn | σTp | σTn | $\mu$ Rpn | $\mu$ Tpn | $\mu$ RTP | $\mu$ RTn |
| --- | --- | --- | --- | --- | --- | --- | --- | --- |
| *1* | 1.58 | 1.59 | 37.48 | 60.39 | 23.77 | 23.62 | 336.25 | 288.85 |
| *2* | **2.16** | **2.38** | **56.72** | 86.59 | 26.70 | **7.77** | 360.74 | 326.27 |
| *3* | **1.78** | 1.58 | 32.35 | 57.59 | 23.25 | **67.43** | 341.00 | **250.32** |
| *4* | 1.59 | 1.47 | 31.63 | 66.97 | 23.91 | **52.34** | 350.72 | 274.47 |
| *5* | 1.36 | 1.54 | **54.18** | **104.01** | 25.59 | 14.32 | **408.53** | **368.62** |
| *6* | 1.69 | 1.69 | 31.81 | 87.27 | 23.05 | 30.60 | 368.83 | 315.18 |

As previously described, specific parameters of TCG varied with ST-T changes associated with ischaemia in PCI. The characteristics of the ECG were explicitly quantified for each parameter.

To compare the TCG results σTn and σTp (red line), μRTp and μRTn (orange line) with the conventional indices of ischaemia ST level and QT interval (blue line), the Mahalanobis distances (MD) during the intervention (Figure 4 B red rectangles C, D, E) were calculated from the distribution during the wait immediately before the intervention (blue rectangles, pr e C, pre D, pre E) as reference, and time-series graphs of the of the MD were obtained (Figures 4C, D, E).

Influences of LCA Angiography (2), the MD of the TCG index increased more than 10 heartbeats earlier than the conventional index, the distance was greater, and the elevation was longer (Figure 4C). In PCI-induced ischaemia, the MD of the TCG index raised more than 20 heartbeats earlier than the conventional index (Figure 4D black arrows). In the late phase of PCI, the TCG index had a greater distance than the conventional index (Figure 4 E). The relatively small transient elevations of the TCG index (red arrows) that were not observed in the conventional index suggested that TCG might be detecting minor ischaemic responses that were not represented in the conventional indexes (Figure 4D, E).

#### Case 2: Early repolarisation syndrome (ERS) presenting ventricular fibrillation

An early repolarisation electrocardiographic pattern is characterised by a sharp, well defined positive deflection or notch immediately following a positive QRS complex at the onset of the ST segment or slurring at the terminal portion of the QRS complex. An imbalance of ionic current between epicardium and endocardium in the ventricle can lead to the development of a transmural voltage gradient that might manifest as an early repolarisation (ER) ECG pattern. An ER-ECG pattern has been confirmed in 6–24% of the general population(17) (18) (19). Prognosis of the majority of the participants with an ER-ECG pattern were considered to be benign. However, it has been reported that a relationship between ER-ECG and sudden cardiac death in a number of the patients showing an ER-ECG pattern. Risk stratification of the ER-ECG pattern is an important issue in clinical practice to prevent sudden cardiac death. Because of the high prevalence of an ER-ECG pattern in the general population, previous st udies showed that it is difficult to distinguish between malignant and benign types of ER-ECG patterns from 12-lead ECG.

A patient with ERS (age in the 20’s) who had a history of multiple repetitive episodes of ventricular tachycardia or ventricular fibrillation (i.e., electrical storm) was tested using TCG.

A subcutaneous implantable cardioverter-defibrillator (ICD) was implanted because of the history of ventricular fibrillation with no structural heart disease. The ECG showed an ER-ECG pattern in inferior-lateral leads and diagnoses as ERS. Frequent episodes of ventricular fibrillation resulted in electrical storm after exchange of the ICD generator.

An ECG (V6) of sinus rhythm 50 pulses prior to ventricular fibrillation (VF) is apparently normal (Figure 5A). The TCG (RT Bulk method) shows that the relative positions of fTn and fTp are reversed and crossed (Figure 5A, black arrow) at the end of the T wave due to a marked elongation of μTn (Figure 5, A black arrow indicating green lines).

**Figure 5.**
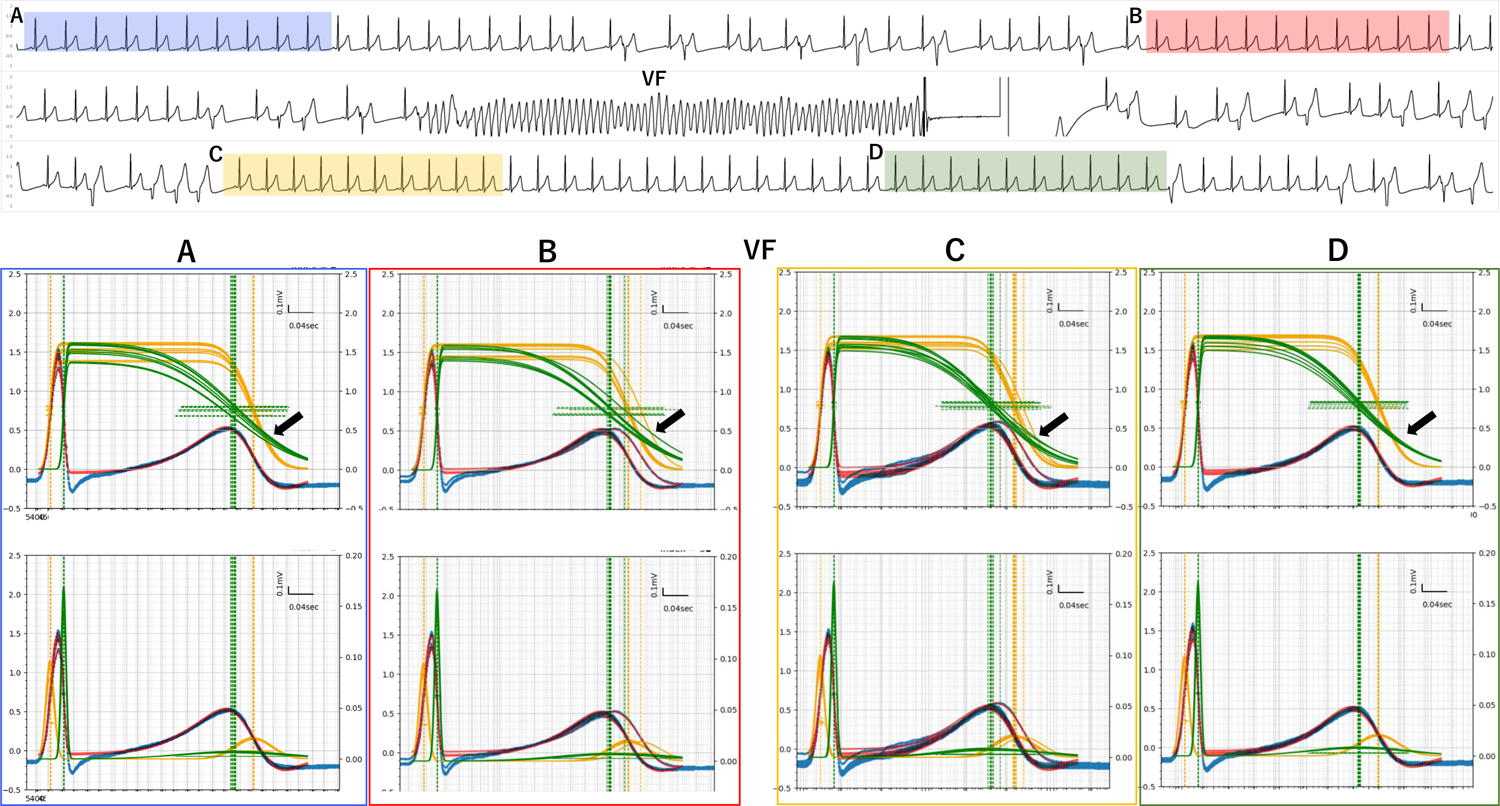
TCG result of malignant early repolarisation syndrome (ERS) presenting ventricular fibrillation (VF) Top ECG (V6) of sinus rhythm 50 pulses prior (**A** blue) 20 pulses before (**B** red) 20 pulses after (**C** yellow) and 45 pulses after (**D** green) VF Bottom 10 pulses overlay of TCG of A∼D sections. Black arrows indicate fTn and fTp reversed and marked elongation and variance of μTn. Red arrows indicate alternans of T waves.

Probability variance of the CDF graph of 10 pulses overlay (Figure 5B) and animation (Supplement Figure 3) represents the marked variance of repolarisation and plateau level. 20 pulses before (B) and after (C) ventricular fibrillation shows alternans of T waves with marked variance of fTn (Figures 5B and C, black arrows indicating green lines) compared with 45 pulses after VF (D).

A crossing of fTn - fTp and fTn - fTp reversal is still present before VF and after returning to sinus rhythm (Figures 5C and D). In TCG parameters, μTn, Tpn and cRTp are significantly increased in ERS (Table 3). This finding is adapted to the characteristic of ERS. Not only is the onset of repolarisation accelerated, but also the delay of repolarisation termination is suggested. Two or three patterns of repolarisation represent the repolarisation complexity of the ER-ECG. TCG can evaluate epicardial and endocardial features in the patients with an ER-ECG pattern to potentially predict high risk patients.

**Table 3.**

| | σRp | σRn | σTp | σTn | μRTp | μRTn | μRpn | μTpn | kRp | kRn | kTp | kTn | $\beta$ | RRI | QT |
| --- | --- | --- | --- | --- | --- | --- | --- | --- | --- | --- | --- | --- | --- | --- | --- |
| A | 6.90 | 3.76 | 28.29 | 89.01 | 325.80 | 266.65 | 22.15 | 36.99 | 1.38 | 1.36 | 1.37 | 1.36 | -0.08 | 820 | 384 |
|  | 6.70 | 3.80 | 29.87 | 86.61 | 324.23 | 271.72 | 21.83 | 30.68 | 1.54 | 1.53 | 1.53 | 1.53 | -0.06 | 822 | 381 |
|  | 6.67 | 3.83 | 30.24 | 86.67 | 324.51 | 273.64 | 21.71 | 29.16 | 1.60 | 1.59 | 1.59 | 1.59 | -0.05 | 819 | 379 |
|  | 6.70 | 3.75 | 30.31 | 85.65 | 324.46 | 272.25 | 21.72 | 30.49 | 1.61 | 1.59 | 1.59 | 1.60 | -0.06 | 822 | 379 |
|  | 6.73 | 3.82 | 30.68 | 85.22 | 324.54 | 274.92 | 21.65 | 27.97 | 1.63 | 1.62 | 1.62 | 1.62 | -0.06 | 820 | 377 |
|  | 6.72 | 3.71 | 29.69 | 87.10 | 325.22 | 270.72 | 21.86 | 32.64 | 1.50 | 1.48 | 1.49 | 1.48 | -0.06 | 820 | 382 |
|  | 6.95 | 3.82 | 29.01 | 88.76 | 325.57 | 266.83 | 22.12 | 36.62 | 1.40 | 1.37 | 1.39 | 1.37 | -0.08 | 816 | 384 |
|  | 6.64 | 3.76 | 29.80 | 86.35 | 324.06 | 268.54 | 21.90 | 33.62 | 1.52 | 1.50 | 1.51 | 1.51 | -0.06 | 818 | 386 |
|  | 6.79 | 3.83 | 30.45 | 85.48 | 324.22 | 272.67 | 21.79 | 29.76 | 1.60 | 1.59 | 1.59 | 1.59 | -0.06 | 819 | 385 |
|  | 6.74 | 3.80 | 30.64 | 85.96 | 324.26 | 272.48 | 21.72 | 30.06 | 1.61 | 1.60 | 1.60 | 1.61 | -0.06 | 819 | 383 |
| B | 6.86 | 3.91 | 32.58 | 87.23 | 345.58 | 297.54 | 21.84 | 26.21 | 1.55 | 1.54 | 1.54 | 1.54 | 0.01 | 804 | 424 |
|  | 6.67 | 3.76 | 31.11 | 89.16 | 327.44 | 276.39 | 21.78 | 29.27 | 1.41 | 1.39 | 1.40 | 1.40 | -0.06 | 806 | 391 |
|  | 6.71 | 3.78 | 30.14 | 87.97 | 326.81 | 273.13 | 21.88 | 31.80 | 1.44 | 1.42 | 1.43 | 1.42 | -0.07 | 809 | 386 |
|  | 6.72 | 3.84 | 31.29 | 85.70 | 325.52 | 276.36 | 21.64 | 27.51 | 1.58 | 1.56 | 1.57 | 1.57 | -0.05 | 811 | 384 |
|  | 6.71 | 3.84 | 31.47 | 85.49 | 325.44 | 276.35 | 21.57 | 27.52 | 1.61 | 1.59 | 1.60 | 1.60 | -0.06 | 815 | 390 |
|  | 6.71 | 3.77 | 31.08 | 85.57 | 325.28 | 276.18 | 21.62 | 27.48 | 1.61 | 1.59 | 1.59 | 1.59 | -0.05 | 819 | 384 |
|  | 6.71 | 3.74 | 30.84 | 83.99 | 325.21 | 274.56 | 21.59 | 29.07 | 1.59 | 1.56 | 1.58 | 1.57 | -0.07 | 824 | 383 |
|  | 6.75 | 3.72 | 29.90 | 88.56 | 326.20 | 272.98 | 21.83 | 31.39 | 1.46 | 1.44 | 1.45 | 1.44 | -0.05 | 829 | 381 |
|  | 6.91 | 3.78 | 29.08 | 87.60 | 326.00 | 270.41 | 21.89 | 33.70 | 1.44 | 1.42 | 1.43 | 1.42 | -0.08 | 829 | 382 |
|  | 6.79 | 3.79 | 30.34 | 85.11 | 325.15 | 274.40 | 21.70 | 29.04 | 1.58 | 1.56 | 1.57 | 1.57 | -0.06 | 834 | 383 |
| C | 7.63 | 4.10 | 31.25 | 82.52 | 324.41 | 265.72 | 21.92 | 36.77 | 1.57 | 1.53 | 1.56 | 1.53 | -0.07 | 741 | 453 |
|  | 7.08 | 3.90 | 29.51 | 78.84 | 310.85 | 249.05 | 21.90 | 39.91 | 1.58 | 1.54 | 1.58 | 1.54 | -0.14 | 736 | 390 |
|  | 7.06 | 3.90 | 30.49 | 78.34 | 309.73 | 251.28 | 21.73 | 36.72 | 1.67 | 1.64 | 1.67 | 1.64 | -0.16 | 734 | 387 |
|  | 6.34 | 3.58 | 31.06 | 92.84 | 312.94 | 254.30 | 21.78 | 36.86 | 1.59 | 1.56 | 1.58 | 1.57 | -0.10 | 733 | 441 |
|  | 6.87 | 3.88 | 30.72 | 78.57 | 309.09 | 250.75 | 21.63 | 36.71 | 1.69 | 1.67 | 1.68 | 1.67 | -0.15 | 731 | 377 |
|  | 6.69 | 3.91 | 30.71 | 74.94 | 307.97 | 246.32 | 21.58 | 40.08 | 1.69 | 1.68 | 1.68 | 1.68 | -0.13 | 731 | 374 |
|  | 6.96 | 3.85 | 30.34 | 78.60 | 309.60 | 250.31 | 21.82 | 37.47 | 1.60 | 1.58 | 1.59 | 1.58 | -0.11 | 734 | 383 |
|  | 7.19 | 3.89 | 30.19 | 79.96 | 310.64 | 251.11 | 21.97 | 37.55 | 1.53 | 1.49 | 1.52 | 1.49 | -0.14 | 735 | 434 |
|  | 7.09 | 3.96 | 30.63 | 76.96 | 309.11 | 250.91 | 21.86 | 36.34 | 1.61 | 1.58 | 1.61 | 1.57 | -0.17 | 736 | 387 |
|  | 6.98 | 3.92 | 31.26 | 78.00 | 309.12 | 253.30 | 21.73 | 34.10 | 1.68 | 1.65 | 1.67 | 1.65 | -0.15 | 738 | 376 |
| D | 6.87 | 3.92 | 31.72 | 77.88 | 309.79 | 256.19 | 21.76 | 31.84 | 1.66 | 1.65 | 1.66 | 1.65 | -0.11 | 750 | 372 |
|  | 6.83 | 3.92 | 31.61 | 77.84 | 309.98 | 256.91 | 21.67 | 31.40 | 1.68 | 1.67 | 1.67 | 1.67 | -0.09 | 749 | 383 |
|  | 6.92 | 3.89 | 31.77 | 76.30 | 309.57 | 256.69 | 21.59 | 31.29 | 1.70 | 1.68 | 1.69 | 1.68 | -0.11 | 751 | 372 |
|  | 6.92 | 3.86 | 31.27 | 78.71 | 310.48 | 256.68 | 21.73 | 32.06 | 1.61 | 1.59 | 1.60 | 1.59 | -0.10 | 753 | 372 |
|  | 7.08 | 3.89 | 30.86 | 80.64 | 311.25 | 255.86 | 21.94 | 33.45 | 1.51 | 1.50 | 1.51 | 1.49 | -0.10 | 757 | 386 |
|  | 6.95 | 3.90 | 30.90 | 79.59 | 311.03 | 255.56 | 21.86 | 33.61 | 1.56 | 1.55 | 1.55 | 1.55 | -0.12 | 757 | 374 |
|  | 6.93 | 3.93 | 31.74 | 76.74 | 310.06 | 258.64 | 21.71 | 29.72 | 1.66 | 1.65 | 1.66 | 1.65 | -0.11 | 758 | 371 |
|  | 6.86 | 3.94 | 31.84 | 77.62 | 310.25 | 258.02 | 21.58 | 30.65 | 1.67 | 1.66 | 1.67 | 1.66 | -0.08 | 759 | 373 |
|  | 6.84 | 3.90 | 31.83 | 78.38 | 310.69 | 259.24 | 21.60 | 29.85 | 1.69 | 1.68 | 1.68 | 1.68 | -0.07 | 760 | 370 |
|  | 6.96 | 3.91 | 32.06 | 77.66 | 310.64 | 259.88 | 21.54 | 29.21 | 1.70 | 1.67 | 1.69 | 1.67 | -0.09 | 763 | 369 |

The cause of the ER-ECG pattern was associated with the transmural voltage gradient between epicardium and endocardium at the early repolarisation phase. An experimental study suggested that heterogeneous loss of the AP dome produces phase 2 re-entry, leading to VF(20). However, the exact mechanism of VF development in patients with ERS was still uncertain. T-wave alternans reflects sudden changes in temporal heterogeneity in ventricular repolarisation and is a sign of cardiac instability leading to life-threatening arrhythmia(21). In addition, microvolt T-wave alternans was confirmed in the patients with ERS(22). In the present case, TCG showed the dynamic nature of the repolarisation alteration mimicking T-wave alternans between epi and endocardial lesions prior to VF development. However, such unique behaviours of the transmural alteration during the repolarisation phase could not be revealed by 12 -lead ECG. TCG can potentially represent the beat-to-beat transmural vulnerability at the repolarisation phase leading to VF.

## Discussion

### ECG model using CDFs: differential of the distribution of myocardial AP transition

A method for estimating the distribution of point processes of collective myocardial AP transitions from the ECG is described. An ECG is generated by a complex system of biological and biophysical mechanisms, we have alleviated the problem of a strict ECG and electromotive force of myocardium relationship and treated the ECG as a point-process model for the timing of the AP transitions and polarity (anode/cathode to ECG) in a myocardium collective.

Differences in myocardial APs, e.g., differences in AP propagation of myocardial fibres in cardiac electrophysiology experiments(23) (10), differences in cross-layer potentials in wedge-shaped myocardium(24) and differences between monophasic APs and location-independent signals in cardiac electrophysiology simulations(14) are closely related to the ECG. Two CDFs were prepared to represent the transition process from resting membrane potential to depolarisation of ventricular muscle as a collective, one representing the transition process of a ventricular muscle collective that is positive to the ECG and the other representing negative. On the basis of their inverse relationship as dipoles, the difference between the two is the ECG equivalent equation, and the parameters of the functions were approximate to those of the ECG. In most case s, the coefficient of the equation determination to the ECG waveform was greater than 0.95.

An approximate equation using the difference between the two CDFs to fit the R and T waves calculates four CDF with metrics mean, standard deviation, weights, and levels. Combining the CDF metrics with the time series of beats and the number of channels in the ECG leads forms a fourth-order tensor. The elements of the tensor and the time-series graph of the four CDFs indicate the variance of collective myocardial AP transitions.

The ECG source, cardiac EMF, is thought to be caused by either 1) the asymmetric shape of heart (hemispheric shape of ventricular wall, without myocardial wall on the basal portion), or 2) asymmetric, complexed patterns of polarisation of the myocardium (depolarisation starts from the endocardial side, propagates to the outer side, after maintaining the depolarised state, repolarisation starts from the outer side and ends on the inner side). If these patterns are normal, the anode positive myocardium group tends to be distributed on the endocardial side and the cathode negative group relatively distributed on the epicardial side. In cardiac physio pharmacology studies, M cells have been proposed as a group of cardiomyocytes with long APDs; the TCG parameters of T positive are thought to correspond to repolarisation of M cells(24)(8).

### AP duration and TCG parameters, μRTp, μRTn

The mean value from the start to end of the μRTp anodic collective AP represents a value close to APD. In contrast, the μRTn cathodic collective AP is shorter than that of APD. The presence of an early repolarising group in the cardiomyocyte collective and a gradual decrease in the plateau potential of the AP to repolarisation may shorten the length of μRTn than that of APD. In fact, a shortening of μRTn was frequently observed in ischaemic myocardium. This is consistent with the shortening or loss of the plateau of the myocardial AP and the rapid descent of the potential after depolarisation (triangular wave) in ischaemic early repolarisation(25) (26).

In the abnormal ECG, such as early repolarisation or repolarisation syndrome as well as in extra systoles and conduction disturbances, the relationship is disrupted. In the clinical ECG cases presented, abnormal patterns of TCG appeared in both positive and negative poles, rather than in the inner-outer (endo-epicardial) relationship.

In particular, the repolarisation-negative component, Tn, tended to be more sensitive to abnormalities instead of localisation, suggesting that it may be useful as a common marker of the abnormalities of cardiac potentials.

As a change in TCG parameters with aging, a prolongation of μRTp and μRTn was observed in the elderly group, suggesting an association with the previously reported elongation of APD in aged cardiomyocytes(27) (28) (29).

### Characteristics of the Bulk and Separation methods

The Bulk method was introduced to improve visibility by connecting the Y-axis of the CDFs and inverse CDFs of the R and T waves in two phases with their heights aligned, creating a trapezoidal shape similar to an AP. As a result, by connecting the anodal and cathodal components of the R and T waves with their respective heights aligned, it was possible to express the relationship of the plateau of AP positive and negative myocardial groups separately. The Bulk method was particularly suitable for the analysis of ischaemic heart disease, where ST changes are important. The Bulk method’s restriction has also been confirmed to increase the stability of the TCG calculation, preventing abnormal results due to fitting errors when ECG contains noise or distortion. However, due to the restriction, it was also confirmed that in the processing of ECGs with complexed abnormalities in waveforms and potential (height), the Bulk method, the fixed effect of the T-wave height (T_k_), may cause distortion and suppression effects on the time axis, such as the mean and standard deviation of the CDF, and may show different results from the Separation method. In such cases, the results of the Separation method were correct. Therefore, it was considered desirable to use both methods depending on the application. In the case studies, the RT Bulk and RT separation methods showed generally similar trends. With the exception of a few abnormal ECGs, the Bulk method is the standard because it provides a clear visual representation of TCG results.

### Relationship between TCG and 12-lead ECG

TCG can be applied to any lead of ECG. Basically, the leads along the main electrical axis of the heart, such as II and CM5, are easy to understand because the direction of depolarisation and repolarisation are straightforward. Also, the thoracic leads from V3 to V6 have a similar relationship due to the proximity of the electrodes to the heart.

TCG results of ECGs recorded from specific electrodes can be analysed in accordance with the corresponding association between electrodes and regions of the heart in conventional ECG interpretation methods. ECG leads, such as AVL V1 V2, are deviated from the major axis of cardiac electrical conduction and the spatial relationship between dipoles during depolarisation and repolarisation is complex; it cannot be simulated with four CDFs and must be represented by additional CDFs, which will be described in the Supplemental section.

### Limitation of TCG Metrics and Myocardial Action Potentials

TCG uses a model that uses a point process in conventional cardiac electrophysiology findings (the dipole model); it is not clear at this time whether the results obtained by TCG are consistent with the actual collective myocardial AP transitions. It should be noted that the plateau phase (Phase 2) of AP is not horizontal and repolarization proceeds slowly in phase 2 with variation between myocytes, which affects the timing of the repolarization transition (Supplement figure 1). As a result, the repolarization timing of the CDF does not always coincide with the acute descending phase (phase3) of the AP waveform in timing. In particular, μTn (the CDF of negative repolarization) thought to be positioned earlier than the previously reported myocardial repolarization timing(8) (24). Further studies using animal experiments and cardiac simulations are needed to confirm the consistency between TCG’s metrics and collective AP transition.

### Application of TCG parameters for Clinical ECG

TCG, which explicitly expresses the relationship between the ECG and collective myocardial AP transition state by introducing statistical measures is described. An application to the clinical ECG suggests that TCG has the potential to capture pathological distortions that are not adequately captured by conventional methods. TCG calculates a time series of three parameters for each of the four CDFs for e ach beat and each lead, and pathological strain is explicitly represented in the tensor. This tensor differs from a physics tensor (such as reciprocity, strain or stress) but rather multi-order data. The information about the temporal and spatial relationships due to the regulated activity of the myocardial collective can be expanded into a definite form as a tensor from the ECG. Using TCG, the probability density distribution (mean μ, variance σ and weight’) of the CDFs can be obtained for the transition process of collective myocardial APs for each beat, making it possible to statistically test for variations and abnormalities in the AP transitions for each beat. For example, the results of TCG for every heartbeat of an individual participant can be statistically tested against the standard values of normal control group.

### Single heartbeat abnormal detection by TCG

It is also possible to test the beat-to-beat statistical difference and/or calculate the mathematical distance between the stable normal condition and unstable abnormal states of the heart disease. The MD can be used as a method for determining the statistical distances. For example, using a certain number of stable-phase TCG results (reference distribution) obtained, the MD from the observed ECG’s TCG results can be determined, which can be used as an index of evaluation, such as abnormal detection.

### Clinical implication of TCG for predicting arrhythmia

Non-sustained ventricular tachycardia (VT) and R on T type of premature ventricular contraction (PVC) are well known as warning arrhythmia that might lead VT/VF3 (30)

However, it is ideal to be able to predict life-threatening arrhythmia by a ECG analysing prior to initiation VT/VF during sinus rhythm. Microvolt T-wave alternans, an oscillation in T-wave morphology, is associated with increased susceptibility to VT/VF (21). Therefore, repolarisation abnormalities are an important sign for the development of VT/VF. The present study showed that TCG could evaluate beat-to-b eat dynamic repolarisation changes between epi and endocardium of the ventricle that did not appear in 12-lead ECG. These results indicated the possibility of the early prediction of life-threatening arrhythmia from a conventional ECG with TCG. Further study will be needed to validate the predictive accuracy of the life-threatening arrhythmia in TCG by prospective study.

## Appendix (Methods)

### Equation for a model of collective ventricular muscle AP transitions by a polar sign-marked point process

The Gaussian distribution of unimodal distribution (normal distribution) representing the probability density of depolarisation time series of the anodic, positive potential (inner layer) of the ventricular muscle, with mean μ_p_ and standard deviation σ_p_ (variance is σ_p_^2^), and the Gaussian distribution of the unimodal distribution representing the probability density of depolarisation time series of the cathodic, negative potential of the ventricular muscle, with mean μ_n_ and standard deviation σ_n_ (variance is σ_p_^2^), are shown in Equations (1) and (2), respectively, where x represents time.

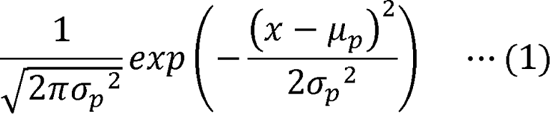

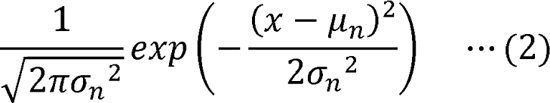

The CDF f_p_(x) in Equation (1) is represented by Equation (3), and the cumulative distribution function (CDF) f_n_(x) in Equation (2) is represented by Equation (4). “erf” is the sigmoid function.

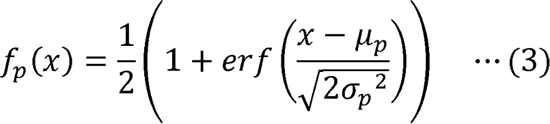

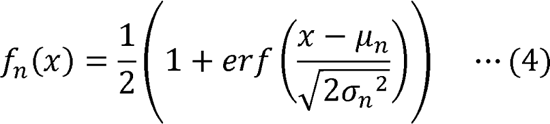

The function of subtracting the second CDF (4) from the first CDF (3) is represented by Equation (5).

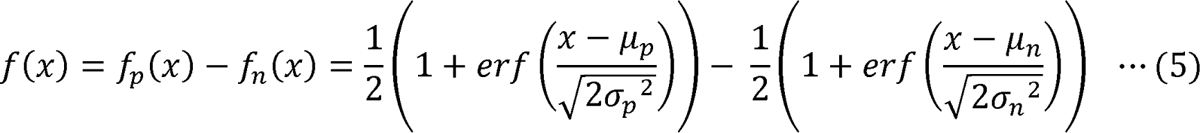

When the CDF *f_p_(x)* in the inner layer and the CDF *f_n_(x)* in the outer layer are weighted and the difference is taken, the weight of *f_p_(x)* is k_p_, the weight of *f_n_(x)* is k_n_ and *f(x)* is expressed by Equation (6).

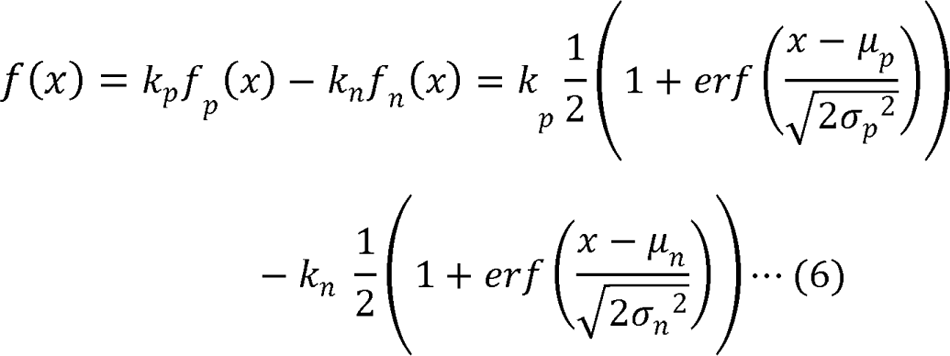

Approximate the target cardiac potential time waveform F(x) with the approximate time waveform f(x), which is the difference between the first and second CDF. The least squares method (L2-norm) can be used for approximation (7).

Least squares fit

*F(x)* Observed ECG data (Target ECG data)

*f(x)* Time sequence probability density function

Minimise the sum of squares of the residuals (L2-norm)

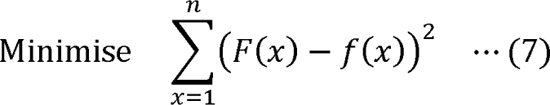

The depolarisation section waveform (R wave) *f_R_(x)* is approximated by the difference between the CDF of the positive and that of the negative. The positive CDF *f_Rp_(x)* is expressed in Equation (8). The negative CDF *f_Rn_(x)* is expressed in Equation (9). The depolarisation interval waveform of the ECG is approximated by the approximate time waveform of Equation (10), which is a function of subtracting the negative CDF *f_Rn_(x)* from the positive CDF *f_Rp_(x)*. The mean μ_Rp_ and standard deviation σ_Rp_, which are parameters that identify the positive CDF, and the mean μ_Rn_ and standard deviation σ_Rn_, which are parameters that identify the negative CDF, are obtained as parameters that represent the characteristics of the depolarisation interval waveform (R wave) (11).

*F_R_(x)* Observed R wave data (Target R wave data)

*f_R_(x)* Time sequence probability density function for R wave

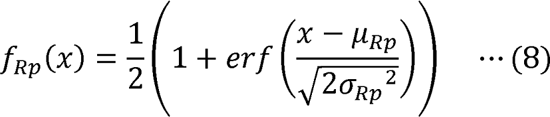

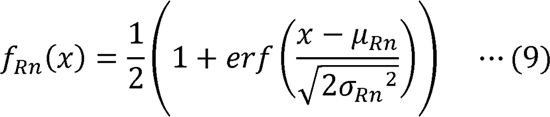

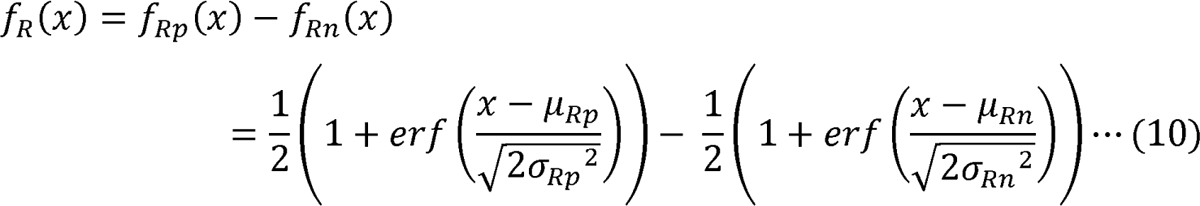

The depolarisation section waveform (i.e., R wave) *f_R_(x)* is differenced by weighting the positive CDF *f_Rp_(x)* and the negative CDF *f_n_(x)*, the weight of *f_Rn_(x)* is k_Rp_ and the weight of *f_n_(x)* is k_Rn_. *F_R_(x)* is expressed in Equation (11).

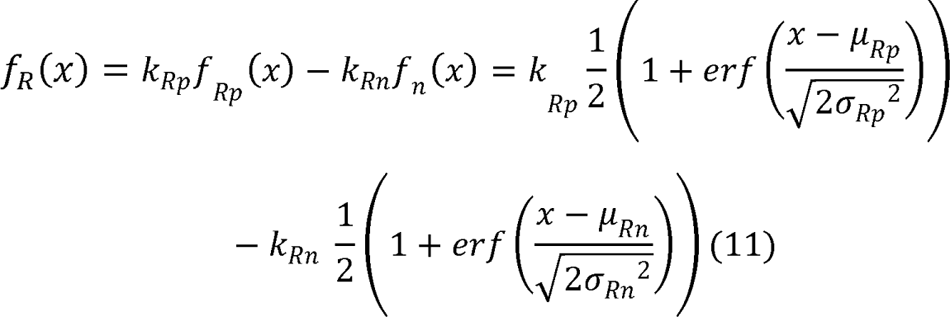

Minimise

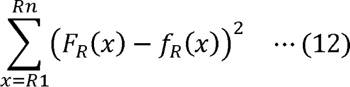

The R wave is a ventricular myocardium sequential depolarisation (OFF to ON), while the T wave is repolarisation (ON to OFF). Thus, the T wave is the opposite switching to the R wave on the time axis.

Therefore, for the repolarisation interval waveform (T wave), two inverse CDF f’_T_(x), where the CDF is subtracted from 1, are used, and the difference or weighted difference between the positive inverse CDF f’_TP_ (x) and the negative inverse CDF f’_Tn_ (x) are approximated by the weighted difference.

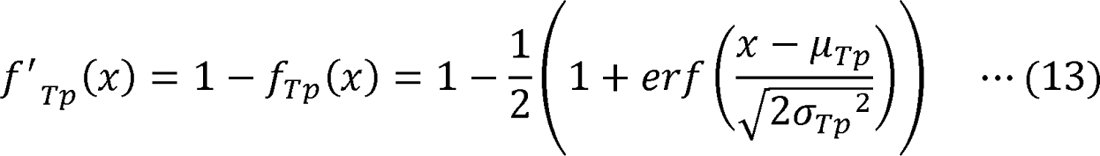

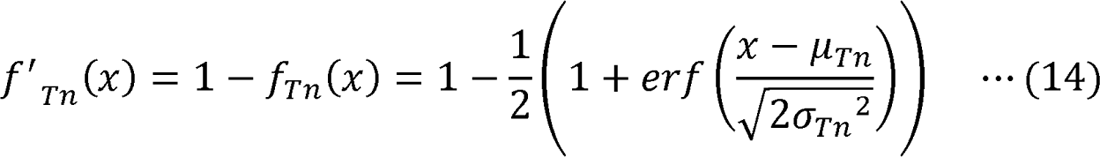

F_T_(x) Observed T wave data (Target T wave data)

f_T_(x) Time sequence probability density function for T wave

f’_T_(x) Reversed time sequence probability density function for T wave

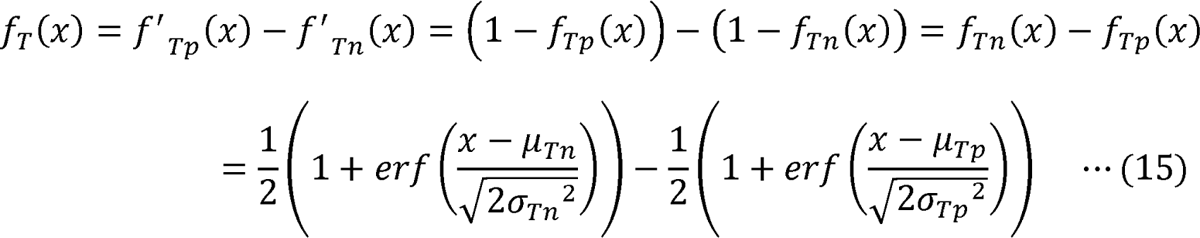

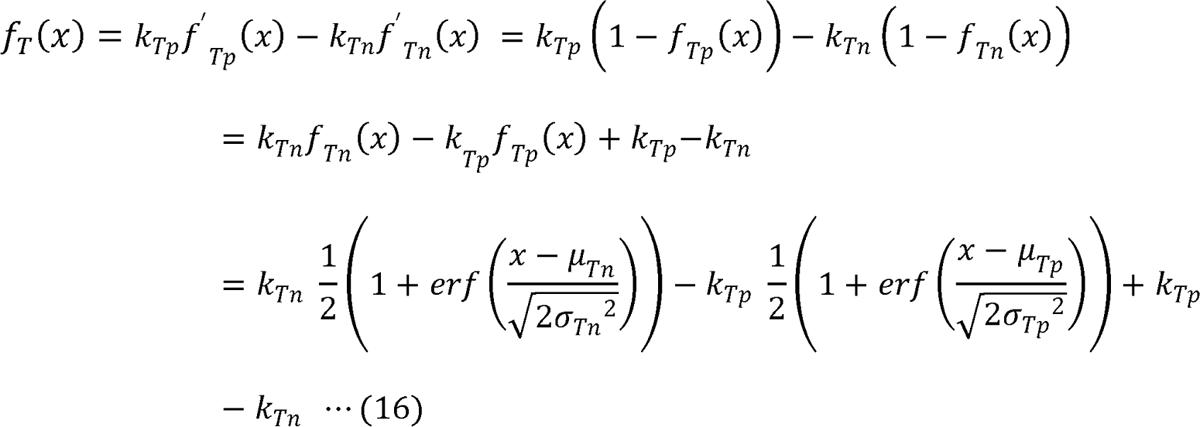

Minimise

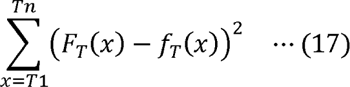

The repolarisation section waveform (T wave) is approximated by the function *f’_Tp_(x)* (inverse positive CDF), which is the subtraction of the positive CDF*f_Tp_(x)* from 1, expressed by Equation (13), and the function *f’_Tn_(x)* (inverse negative CDF), which is the subtraction of the negative CDF *f_Tn_(x)* from 1, expressed in Equation (14).The repolarisation time waveform is approximated by Equation (15), which is a function of subtracting the inverse negative CDF *f’_Tn_(x)* from the inverse positive CDF *f’_Tp_(x)s*.

When the repolarisation interval waveform (T wave) is approximated by the weighted difference between the inverse positive CDF *f’_Tp_(x)* and inverse negative CDF *f’_Tn_(x)*, the repolarisation time waveform is approximated by the approximate time waveform of Equation (16), which is the function of multiplying the inverse positive CDF *f’_Tp_(x)* by the weight k_Tp_ and subtracting the function of multiplying the inverse negative CDF *f’_Tn_(x)* by the weight k_Tn_.

The difference between the observed ECG R wave and the CDF*f_Rp_(x)-f’_Rn_(x)*, and the difference between the T wave and *fs_Tn_(x)-f’_Tp_(x)* are minimised, and the parameters of the four CDFs are obtained.

The weights k of the CDFs of depolarisation (R wave) and repolarisation (T wave) are determined by two methods: 1) the method of determining the four independent parameters of the positive and negative (RT Separation method, Figure 2A) the method of determining k under the condition that weights k of the depolarisation (R wave) and repolarisation (T wave) CDFs are equal, which means the positive pair *f_Rp_(x),f’_Tp_(x)* and negative pair *f_Rn_(x), f’_Tp_(x)* are connected in the plateau (RT Bulk method, Figure 2B).

(RT Bulk method, the points between the R peak and the R peak + 60 msec that are lower than the ST junction point are excluded from the fitting. This is to prevent the s downward peak from adversely affecting the fitting.)

The computer program calculated all four CDFs parameters and the CDFs intervals as well as conventional ECG variables including RR interval, QT time, and ST level.

The μRTp, the interval between μRp to μTp, represents the average duration of positive collective APs, and μRTn, μRn to μTn represents the duration of negat ive collective APs (Figure 2B).

The results of this analysis can be expressed as a matrix of in fourth order tensor, three dimensions (ECG leads, CDFs and each parameters) and one dimension in time. For example, the number of ECG leads (e.g., I, II, V2), CDFs, the parameters of distribution (Average μ, Standard deviation σ, Weight k, baseline level β) and time of heartbeat <3,4,4,1> (Supplement Figure 3).

### Extensions to the complexed ECG

ECGs with abnormal waves (e.g., delta and J waves), T-wave diversity (e.g., with inflection points) and the complicated excitation propagation projected leads (e.g., aVL, V1, V2) cannot be represented completely with four CDFs. In such cases, CDFs are added to increase the expressiveness. For example, the extra CDF is able to add. Specifically, abnormal waves that appear around R and T waves, such as delta and J waves, or with irregularities or inflection points (subtle wave) not normally seen in ST and T waves, can be handled by adapting the same process previously described for R and T waves to abnormal waves. For example, when approximating Δ waves *F_D_* which appear between R and T waves, the *f_D_* difference between the CDF f_DP_(x) and CDF *f_Dn_(x)* is approximated by the time waveform of Equation (18), and the abnormal waveforms are obtained as parameters that represent the characteristics of the waves.

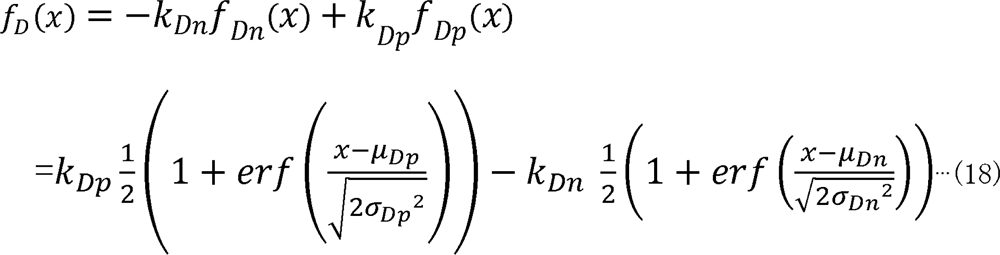

Minimise

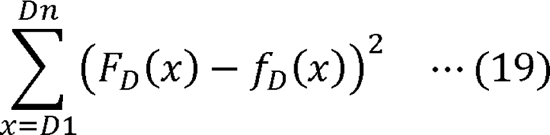

## Data Availability

We are currently consulting with the Ethics Committee of Nippon Medical School regarding the disclosure of ECG and TCG data included in the case reports (PCI and ERS?. If approved, the corresponding author will provide the data upon request.

---

**Supplement Figure 1.**
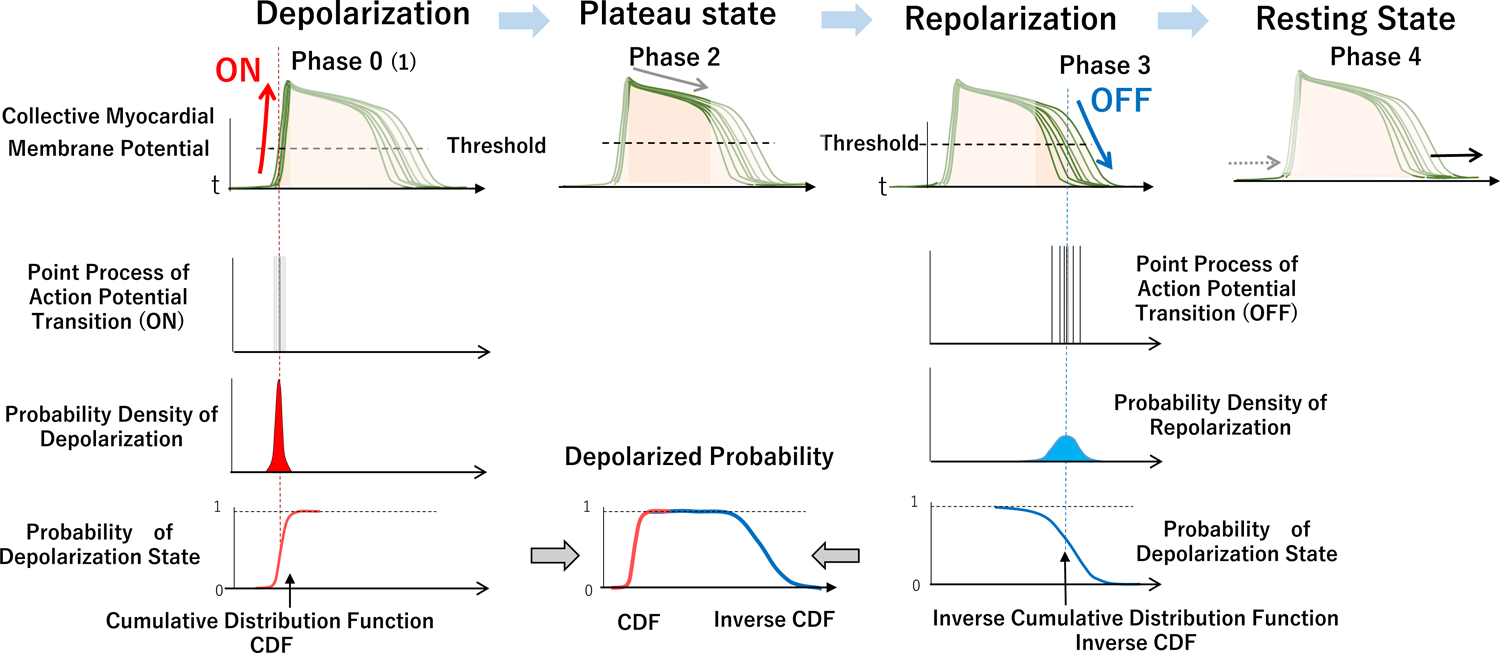
Graphs and point processes expression of collective myocytes of AP transitions, probability density distribution and CDF. Depolarisation process is synchronous and the transitions are concentrated in short periods. The potential of myocytes repolarises after a plateau, and the transition process is distributed. The probability of depolarisation is expressed by a CDF (red) and inverse CDF (blue).

**Supplement Figure 2.**
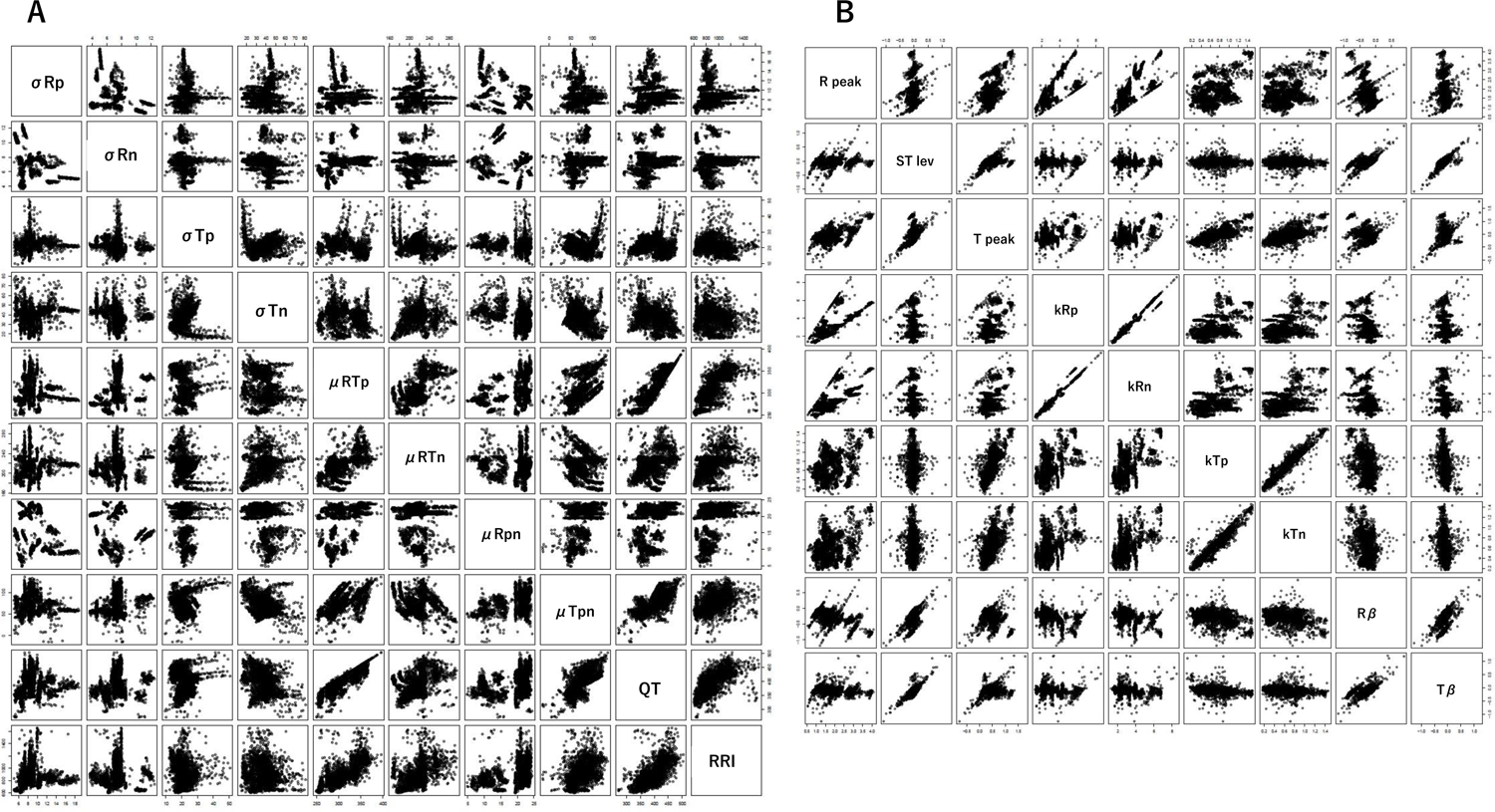
Scatter plot of Supplement Table 1A (**A**) and B (**B**)

**Supplement Figure 3.**
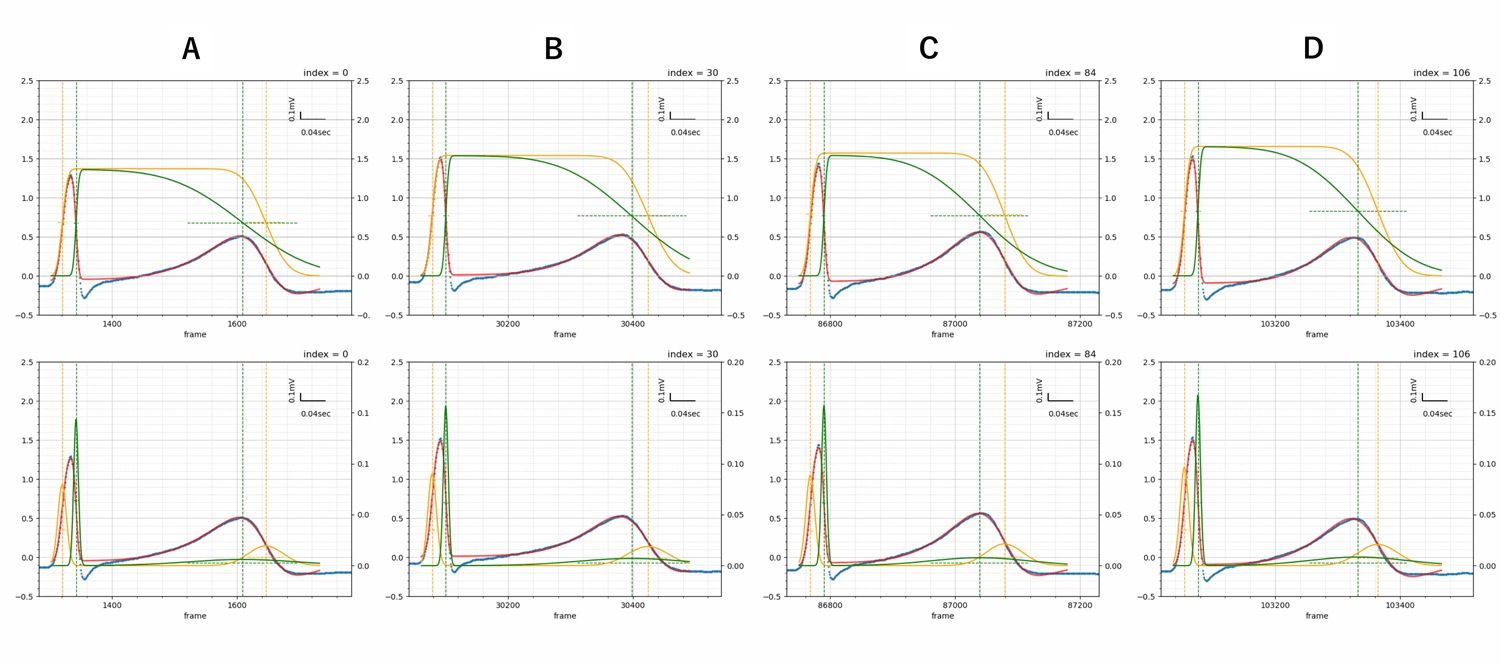
TCG of continuous sinus rhythm 10 beats of GIF animation (Figusres 5A∼D)

**Supplement Figure 4.**
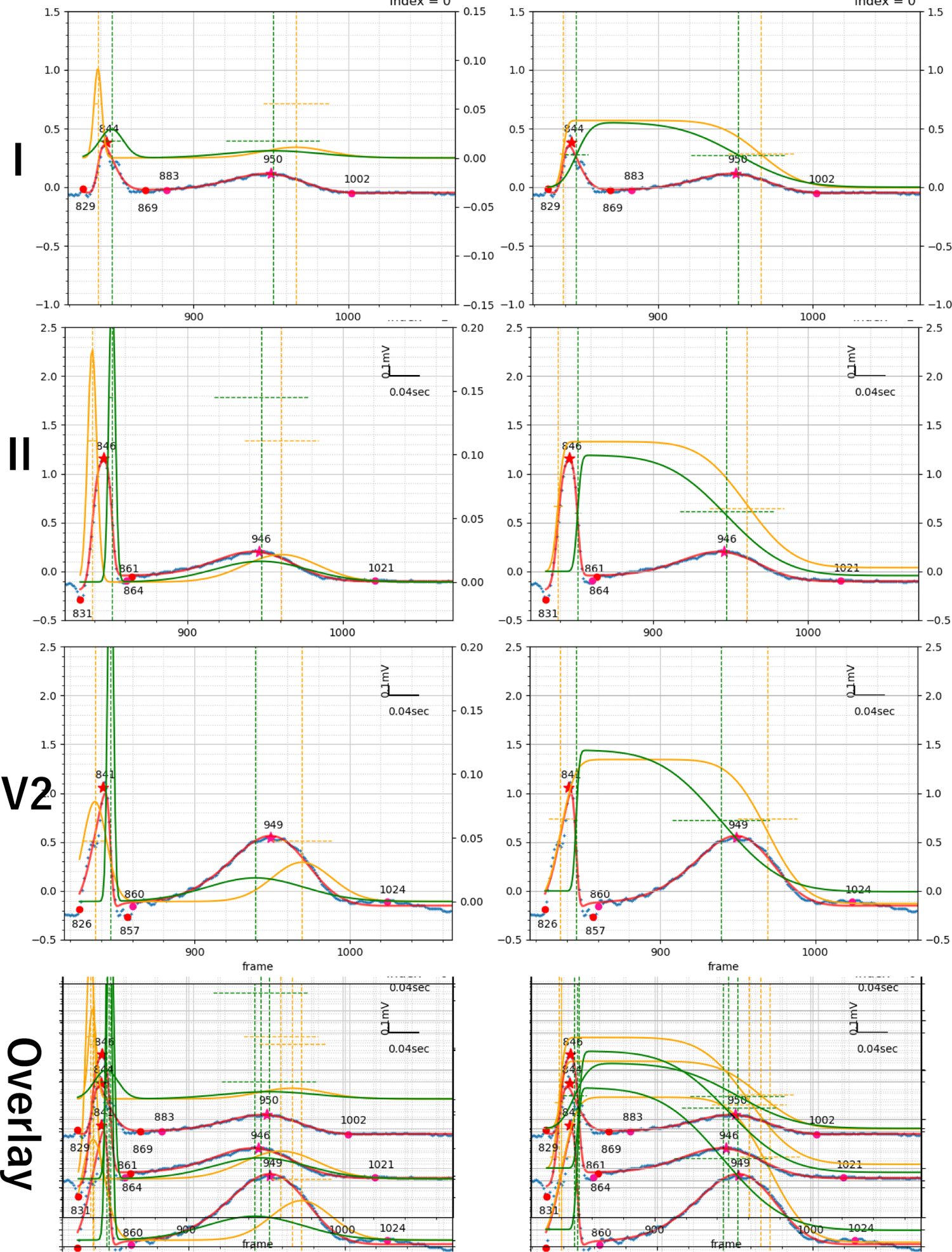
TCG (Bulk method) result of I, II, V2, of standard 12 leads ECG

## Authors’ contributions

Shingo Tsukada created the conception of TCG, designed the statistical equation model, adapted it to ECG data, performed the data processing and discussed based on electrophysiology. Yu-ki Iwasaki and Yayoi Tetsuo Tsukada contributed data collection of clinical electrocardiogram, assisted in text and figure corrections of the manuscript, supervised clinical implication of the study.

## Acknowledgements

The authors would like to thank A. Shiozawa (NTT DATA Mathematical Systems Inc.) for computer coding of TCG. This study was supported by KAKENHI Grants-in-Aid for Scientific Research (22K08217) and NTT’s internal basic research grant (IOWN). All authors were not precluded from accessing data in the study, and accepted responsibility to submit for publication.

## Ethics committee approval

This study was approved by the Nippon Medical School Ethics Committee.

---

**Supplement Table 1.**

| A |  |  |  |  |  |  |  |  |  |  |
| --- | --- | --- | --- | --- | --- | --- | --- | --- | --- | --- |
| | $\sigma_{Rp}$ | $\sigma_{Rn}$ | $\sigma_{Tp}$ | $\sigma_{Tn}$ | $\mu_{RTp}$ | $\mu_{RTn}$ | $\mu_{Rpn}$ | $\mu_{Tpn}$ | QT | RRI |
| $\sigma_{Rp}$ | | -0.34 | 0.09 | 0.14 | -0.11 | 0.04 | -0.41 | -0.10 | -0.02 | -0.03 |
| $\sigma_{Rn}$ | -0.34 | | -0.07 | -0.12 | 0.26 | 0.11 | 0.14 | 0.23 | 0.19 | 0.04 |
| $\sigma_{Tp}$ | 0.09 | -0.07 | | 0.02 | -0.14 | -0.25 | -0.11 | 0.09 | 0.04 | -0.13 |
| $\sigma_{Tn}$ | 0.14 | -0.12 | 0.02 | | -0.16 | 0.28 | -0.33 | -0.44 | -0.33 | -0.17 |
| $\mu_{RTp}$ | -0.11 | 0.26 | -0.14 | -0.16 | | **0.66** | 0.33 | **0.67** | **0.88** | **0.73** |
| $\mu_{RTn}$ | 0.04 | 0.11 | -0.25 | 0.28 | 0.66 | | -0.04 | -0.10 | 0.42 | 0.48 |
| $\mu_{Rpn}$ | -0.41 | 0.14 | -0.11 | -0.33 | 0.33 | -0.04 | | 0.27 | 0.35 | 0.29 |
| $\mu_{Tpn}$ | -0.10 | 0.23 | 0.09 | -0.44 | 0.67 | -0.10 | 0.27 | | **0.74** | 0.47 |
| QT | -0.02 | 0.19 | 0.04 | -0.33 | 0.88 | 0.42 | 0.35 | 0.74 |  | **0.66** |
| RRI | -0.03 | 0.04 | -0.13 | -0.17 | 0.73 | 0.48 | 0.29 | 0.47 | 0.66 |  |

| B |  |  |  |  |  |  |  |  |  |
| --- | --- | --- | --- | --- | --- | --- | --- | --- | --- |
|  | R peak | ST lev | T peak | kRp | kRn | kTp | kTn | Rβ | Tβ |
| R peak |  | 0.12 | 0.54 | **0.74** | **0.76** | 0.40 | 0.33 | -0.47 | -0.08 |
| ST lev | 0.12 |  | 0.49 | -0.01 | -0.02 | 0.05 | 0.00 | 0.56 | **0.80** |
| T peak | 0.54 | 0.49 |  | 0.43 | 0.44 | **0.74** | **0.67** | -0.13 | 0.17 |
| kRp | 0.74 | -0.01 | 0.43 |  | **0.98** | 0.46 | 0.44 | -0.37 | -0.12 |
| kRn | 0.76 | -0.02 | 0.44 | 0.98 |  | 0.46 | 0.44 | -0.35 | -0.13 |
| kTp | 0.40 | 0.05 | 0.74 | 0.46 | 0.46 |  | **0.95** | -0.41 | -0.29 |
| kTn | 0.33 | 0.00 | 0.67 | 0.44 | 0.44 | 0.95 |  | -0.39 | -0.17 |
| Rβ | -0.47 | 0.56 | -0.13 | -0.37 | -0.35 | -0.41 | -0.39 |  | **0.65** |
| Tβ | -0.08 | 0.80 | 0.17 | -0.12 | -0.13 | -0.29 | -0.17 | 0.65 |  |

**Supplement Table 2.**
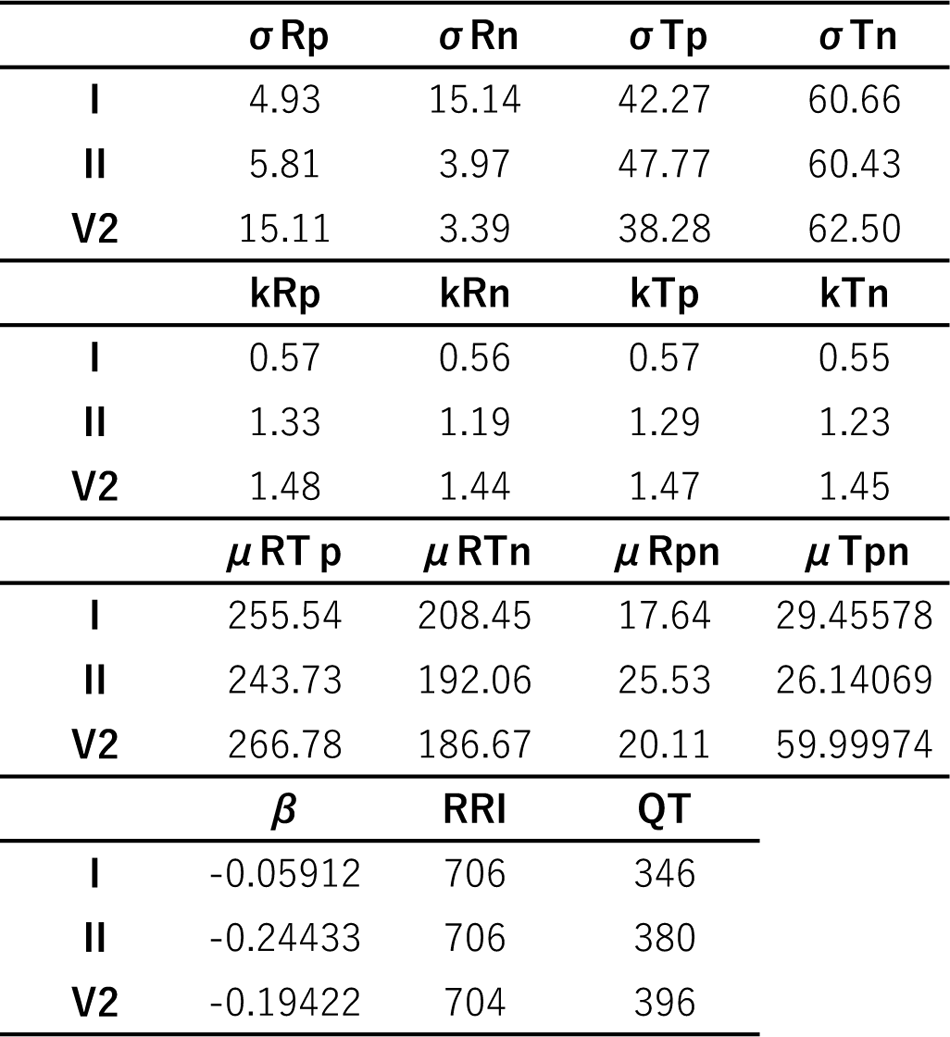

